# Biomarker Discovery in Alzheimer’s and Neurodegenerative Diseases using Nucleic Acid-Linked Immuno-Sandwich Assay

**DOI:** 10.1101/2024.07.29.24311079

**Authors:** Nicholas J. Ashton, Andrea L. Benedet, Guglielmo Di Molfetta, Ilaria Pola, Federica Anastasi, Aida Fernández-Lebrero, Albert Puig-Pijoan, Ashvini Keshavan, Jonathan Schott, Kubra Tan, Laia Montoliu-Gaya, Richard Isaacson, Matilde Bongianni, Chiara Tolassi, Valentina Cantoni, Antonella Alberici, Alessandro Padovani, Gianluigi Zanusso, Andrea Pilotto, Barbara Borroni, Marc Suárez-Calvet, Kaj Blennow, Henrik Zetterberg

## Abstract

**INTRODUCTION:** Recent advancements in immunological methods accurately quantify biofluid biomarkers for identifying Alzheimer’s pathology and neurodegeneration. Despite this progress, more biomarkers, ideally in blood, are needed for effective patient management and disease monitoring for Alzheimer’s disease (AD) and other neurodegenerative proteinopathies.

**METHODS:** We employed the Nucleic Acid-Linked Immuno-Sandwich Assay (NULISA™) central nervous system (CNS) panel for biomarker quantification in plasma, serum and cerebrospinal fluid (CSF) of patients with AD, mild cognitive impairment, Lewy body dementia, progranulin (*GRN*) mutation carriers and matched controls.

**RESULTS:** NULISA™ identified p-tau217 and NfL as the most significantly deregulated plasma biomarkers in the AD continuum and *GRN* mutation carriers, respectively. Importantly, numerous novel and significant proteomic changes were observed in each disease comparison, which included proteins involved in synaptic processing, inflammation, microglial reactivity, TDP-43 and α-synuclein pathology. Plasma and serum act as complimentary biofluids.

**CONCLUSION:** We highlight the potential of next-generation biomarker identification tools, such as NULISA™, to detect novel proteomic features that incorporate established biomarkers like p-tau217 and NfL. These findings highlight the importance of continued biomarker discovery to enhance patient management, improve treatment decisions, and better understand the complexities of neurodegenerative disorders.

## INTRODUCTION

Discovery plasma proteomics has long since demonstrated a differential signal in the blood of patients clinically diagnosed with Alzheimer’s disease (AD) ^1,2^. These earlier studies used agnostic methods to discover novel biomarkers—proteomic technologies not predicated on any *a priori* hypotheses in a case-control approach ^1,3–6^ but later evolved to an endophenotype design based on pathology; brain atrophy measures ^1,7–9^, cerebrospinal fluid (CSF) biomarkers ^10^, or amyloid-β (Aβ) positron emission tomography (PET) ^11–15^. These reports continually implicated a dysregulated complement, coagulation, acute inflammation response ^5,16^ and markers previously identified in genome-wide association studies (e.g., clusterin ^17^) but failed in identifying a replicable biomarker signature with the required disease specificity or clinical usefulness.

The advancement in ultra-sensitive immunoassays and highly specific immunoprecipitation mass spectrometry (IP-MS) methods that are complemented by an increasing number of well-characterised research cohorts, adhering to a biological definition of disease ^18^, have substantially contributed to the rapid development of key dementia targets in blood samples. In Alzheimer’s, measures of phosphorylated tau (p-tau), particularly p-tau217 ^19,20^, have shown utility in identifying cerebral pathology at all stages of the disease *continuum*. Thus, it is anticipated that plasma p-tau217 will be an essential component of patient management and treatments decisions related to AD. As a disease-defining biomarker in CSF, the detection of Aβ42/40) in plasma has seen much improvement ^21,22^, and while it is unlikely to feature as a primary diagnostic blood biomarker, due to several confounding factors (*e.g.,* poor robustness ^23^, pharmacodynamic effects ^24^ and pre-analytical instability), it will have relevance in understanding novel disease-modifying therapies (DMT) that shift Aβ production to a less pathogenic state (*e.g.,* gamma-secretase modulators).

Yet, there remains a multitude of biomarker applications in AD and related disorders where plasma p-tau217 and Aβ42/40 are not fully sufficient. These blood biomarkers do not entirely explain the extend of tau accumulation ^25^, longitudinal clinical progression ^26^, or response to amyloid clearance ^27^, although p-tau217 is the leading candidate of such associations. In the light of approved DMT, prognostic biomarker signatures of responders and non-responders to interventions or those at risk of adverse events are greatly needed to guide treatment management prior to administration. Further, p-tau217 and Aβ42/40 only work to identify Alzheimer pathology as the main or contributing factor in other neurogenerative proteinopathies. AD accounts for ∼60% of all diagnosed dementia cases, and supplemental biomarkers are required to differentiate other forms of dementia to further improve patient management and accelerate drug development. Promising results from α-synuclein (αSyn) seed amplification assays (SAA) in Lewy body (LB) dementia have not yet translated from CSF to blood and biomarkers of vascular, TDP-43 and 4R tau pathologies remain unresolved. In blood, neurofilament light (NfL) shows limited disease specificity ^28^, with some isolated instances *e.g.,* atypical parkinsonian disorders compared to Parkinson’s disease ^29^ but remains a useful biomarker for general neurodegeneration and acute neurological injury. Glial fibrillary acidic protein (GFAP) in blood has also shown changes in some non-Alzheimer pathologies ^30,31^ but recent data has clearly demonstrated its dynamic change in response to amyloid and tau deposition ^32–35^. Thus, direct, or indirect, measures in blood that are more specific to other proteinopathies are urgently needed.

Proteomic studies of biofluid biomarkers can be targeted or non-targeted; the latter often guiding the former. A common non-targeted approach to identify blood-based biomarkers is liquid chromatography– tandem mass spectrometry (LC–MS/MS) but has limited coverage of the plasma proteome. This is due to the high dynamic range of protein abundances in blood, untargeted LC-MS/MS has the limitation that signals derived from low-abundant proteins are often masked by those of higher concentrations, thus making LC-MS/MS not optimal for the detection of those small changes, particularly at the sub-picomolar level ^11,12^, Instead, high plex solutions with attomolar sensitivity, such as Olink, SomaLogic, and now NULISA™, can offer targeted methods with *hundreds-to-thousands* of proteins previously implicated in disease being screened in low volume and simplified workflows.

This pilot study does not aim to resolve the unmet needs highlighted above such as the identification of novel biomarkers, but it aims to provide an example of a new direction in Dementia proteomic technologies which combines robust single analytes of neurodegenerative disease within multi-analyte and high-throughput panel. This study utilises the Nucleic Acid-Linked Immuno-Sandwich Assay (NULISA™) CNS panel in four cohorts with different biomarkers profiles; control (Aβ-) and AD (Aβ+), mild cognitive impairment (MCI Aβ+/-), Lewy body pathology (LB+/-) and progranulin (*GRN*) carriers and non-carriers.

## METHODS

### Participants & Ethics

In cohort 1, forty participants were collected from the prospective University College London Dementia Research Centre as a part of Alzheimer’s Association Global Biomarker Standardisation Consortium (GBSC) plasma phospho-tau Round Robin study. Participants were divided based on their CSF Aβ42/40 and p-tau181 (Lumipulse G; Fujirebio, Belgium) concentrations. Participants were confirmed as AD pathology if CSF Aβ42/40 was <0.065 and p-tau181 was >57 pg/ml. All participants gave written informed consent according to the Declaration of Helsinki. The study was approved by the local Ethics Committee (Wolfson CSF study 12/0344). In cohort 2, forty participants were enrolled from the BIODEGMAR cohort, an observational longitudinal study that enrols individuals with cognitive symptoms and/or neurodegenerative diseases visiting the Cognitive and Behavioural Unit of Hospital del Mar (Barcelona, Spain)^36^. Core AD CSF biomarkers (Aβ42/40, p-tau, and t-tau) were measured with Lumipulse immunoassays (Fujirebio, Belgium). Participants were classified as AD CSF profile if the CSF Aβ42/p-tau ratio was <10.25^36^. In cohort 3, fifty-four participants were enrolled at the Neurology Unit of Brescia. The presence or absence of Lewy Body (LB+/-) pathology was determined by α-synuclein (αSyn) seed amplification assay (SAA) at the university of Verona as previously reported^37^. The LB-patients met the NIA-AA clinical criteria for the diagnosis of AD (n=30) confirmed based on a CSF biomarker profile (p-tau/ Aβ42 ratio > 0.9) ^38^. The LB+ patients were clinically classified as DLB (n=22), according to the definitions and guidelines provided by the DLB Consortium, and PD (n=2) according to current clinical criteria^39^. All participants gave written informed consent according to the Declaration of Helsinki. The study was approved by the local Ethics Committee (NP 1471). In cohort 4, forty participants were enrolled at the Center for Neurodegenerative Disorders, University of Brescia (Italy) from October 3rd, 2023, to November 23rd, 2023. In FTD patients (n=18), the presence pathogenetic *GRN* mutations was carried out according to standard procedures ^40^. Presymptomatic *GRN* mutation carriers (n=2) ^41^ and healthy controls (HC, n=20), recruited among spouses or patients’ family members, were also included. The FTD patients met current clinical criteria for the diagnosis of agrammatic variant of primary progressive aphasia (avPPA, n=11), behavioural variant frontotemporal dementia (bvFTD, n=6), semantic variant of primary progressive aphasia (svPPA, n=1). All participants gave written informed consent according to the Declaration of Helsinki. The study was approved by the local Ethics Committee (NP 2189). A summary of patient demographic and clinical characteristics for all four cohorts are shown in **Table 1**.

**Table 1.** The demographic information for cohorts 1-4.

|  | *N* | Age,<br>mean<br>(SD) | Sex, %F | APOE e4<br>carriership, % | Biomarker |
| --- | --- | --- | --- | --- | --- |
| **Cohort 1** |  |  |  |  |  |
| Non-AD | 15 | 63.1 (5.2) | 66.7% | 26.7% | 0.093 (0.022) <sup>a</sup> |
| AD | 25 | 65.2 (3.4) | 64% | 72% | 0.045 (0.012) <sup>a</sup> |
| **Cohort 2** |  |  |  |  |  |
| MCI+ | 25 | 73.8 (6.2) | 64% | 13.3% | 4,9 (5,1) <sup>b</sup> |
| MCI- | 15 | 73.4 (4,6) | 60% | 56% | 23,8 (7,1) <sup>b</sup> |
| **Cohort 3** |  |  |  |  |  |
| AD | 30 | 77.2 (7.1) | 50% | 66.7% | 1,02 (0.07) <sup>d</sup><br>0% <sup>e</sup> |
| LB | 24 <sup>c</sup> | 69.3 (4.1) | 41.6% | 25% | 0,74 (0.51) <sup>d</sup><br>100% <sup>e</sup> |
| **Cohort 4** |  |  |  |  |  |
| GRN- | 20 | 65.4 (5.2) | 65% | NA | 20 |
| GRN+ | 20 <sup>f</sup> | 63.1 (4.7) | 65% | NA | 20 |

<sup>a</sup> CSF Aβ42/40 <0.065; <sup>b</sup> CSF Aβ42/p-tau ratio <10.25; <sup>c</sup> Dementia with Lewy Body, 22; Parkinson's disease, 2; <sup>d</sup> p-tau/ β42 ratio > 0.9; <sup>e</sup> SAA+; <sup>f</sup> avPPA, 11; bvFTD, 6; svPPA, 1; pre-symptomatic, 2

### NUcleic acid Linked Immuno-Sandwich Assay (NULISA^TM^) analysis

NULISA^TM^ assays were performed at Alamar Biosciences as described previously ^42^. Briefly, plasma, serum, and CSF samples stored at -80°C were thawed on ice and centrifuged at 10,000*g* for 10mins. Exactly 10 μL supernatant were plated in 96-well plates and analyzed with Alamar’s CNS Disease Panel targeting mostly neurodegenerative disease related targets as well as inflammation and immune response-related cytokines and chemokines. A Hamilton-based automation instrument was used to perform the NULISAseq workflow. Briefly, immunocomplexes were formed with DNA-barcoded capture and detection antibodies, followed by a capturing and washing step of the immunocomplexes on paramagnetic oligo-dT beads. The immunocomplexes were then released into a low-salt buffer, which were captured and washed on streptavidin beads. Finally, the proximal ends of the DNA strands on each immunocomplex were ligated to generate a DNA reporter molecule containing both target-specific and sample-specific barcodes. DNA reporter molecules were pooled and amplified by PCR, purified and sequenced on Illumina NextSeq 2000. Singleplex NULISA assays were performed by Alamar Bioscience on the Argo prototype instrument. For the p-tau217 assay, 20 µL of plasma or CSF were used in a reaction volume of 100 µL containing capture and detection antibody cocktails in Assay Diluent buffer. The mixture was incubated at room temperature for 1-hour to allow the formation of immunocomplexes. These immunocomplexes then underwent two rounds of affinity bead purification and multiple washes, followed by ligation to form reporter molecules, which were then eluted. The final eluate was collected for quantification by qPCR.

### Data processing and normalization

For NULISAseq, sequencing data were processed using the NULISAseq algorithm (Alamar Biosciences). The sample- (SMI) and target-specific (TMI) barcodes were quantified, and up to two mismatching bases or one indel and one mismatch were allowed. Intraplate normalization was performed by dividing the target counts for each sample well by that well’s internal control counts. Interplate normalization was then performed using interplate control (IPC) normalization, wherein counts were divided by target-specific medians of the three IPC wells on that plate. Data were then rescaled, add 1 and log2 transformed to obtain NULISA Protein Quantification (NPQ) units for downstream statistical analysis. For singleplex assays, the Cq read out from the calibrators and samples was transformed to a linear scale using the formula 2^(37−Cq). The linear transformed signal was then used in the 4-parameter logistic (4-PL) curve fitting for standard curve generation and sample analyte concentration calculation.

### Statistical analysis

All statistical analyses and plots were performed on R statistical software version 4.4.0. For descriptive statistics chi-square tests compared categorical variables between groups and one-way analysis of variance (ANOVA) compared continuous variables between groups. Spearman rank tests (confidence intervals (CI) at 95% were determined with 1000 bootstraps resampling) tested for correlations between biomarker assays. Mean-fold changes were computed using the average of the respective control group for each analysis. The area under the receiver operating characteristic curve (AUROC) assessed the biomarker accuracy to distinguish predefined statuses and 95% CI of sensitivities and specificities were also computed (Youden index). Linear Models for Microarray and RNA-Seq Data (LIMMA models) evaluated the differential protein expression between designated groups. Volcano plots report by color schemes the proteins that were nominally significant as well as the proteins that remained significant after false discovery rate (FDR) correction for multiple comparisons.

## RESULTS

This study included four pilot datasets totalling 190 participants (**Table 1**). In details, 40 participants (mean [SD] age, 63.8 [5.9] years; n [%] 17 females [42.5%]) biologically determined as AD or non-AD by CSF biomarkers (cohort 1), from the Alzheimer’s Association Global Biomarker Standardisation Consortium (GBSC) plasma phospho-tau Round Robin study; 40 participants (mean [SD] age, 73.7 [5.9] years; n [%] 25 females [62.5%]) clinically determined identified as MCI with or without AD pathology determined by CSF Aβ42/p-tau181 (cohort 2); 54 participants (mean [SD] age, 76.3 [7.8] years; n [%] 24 females [44.4%]) with or without Lewy body pathology by CSF biomarkers (cohort 3) and 40 participants (mean [SD] age, 63.9 [6.9] years; n [%] 26 females [65.0%]) with (n=20) or without a *GRN* mutation (cohort 4).

### NULISAseq in biomarker-defined AD compared to non-AD

In the plasma of biologically determined AD compared to non-AD (cohort 1), we observed six differentially deregulated proteins (**Fig. 1A**), of which, four upregulated proteins in AD passed multiple testing correction (p-tau217, logFC = 1.68, *P_adj_* < 0.001; GFAP, logFC = 0.90, *P_adj_* = 0.003; p-tau231, logFC = 0.90, *P_adj_* = 0.003; BACE1, logFC = 0.33, *P_adj_* = 0.02 **Suppl. Fig. 1**). The boxplots of nominally significant proteins are shown in **Suppl. Fig. 2**. Interestingly, in the serum of the same patients, we observed 18 significantly changed proteins (**Fig. 1B**) of which two upregulated proteins passed multiple testing correction (p-tau217, logFC = 1.33, *P_adj_*< 0.001; GFAP, logFC = 1.02, *P_adj_* < 0.001; **Suppl. Fig. 3**). The boxplots of nominally significant proteins are shown in **Suppl. Fig. 4**. In the CSF of the same patients, 22 proteins significantly changed were observed (**Fig. 1C**), of which, one downregulated and 14 upregulated proteins passed multiple testing correction (**Suppl. Fig. 5**). The boxplots of nominally significant proteins are shown in **Suppl. Fig. 6.** The results for all proteins measured in the NULISAseq CNS panel for plasma, serum and CSF are displayed in **Suppl. Tables 1-3**.

**Figure 1.**
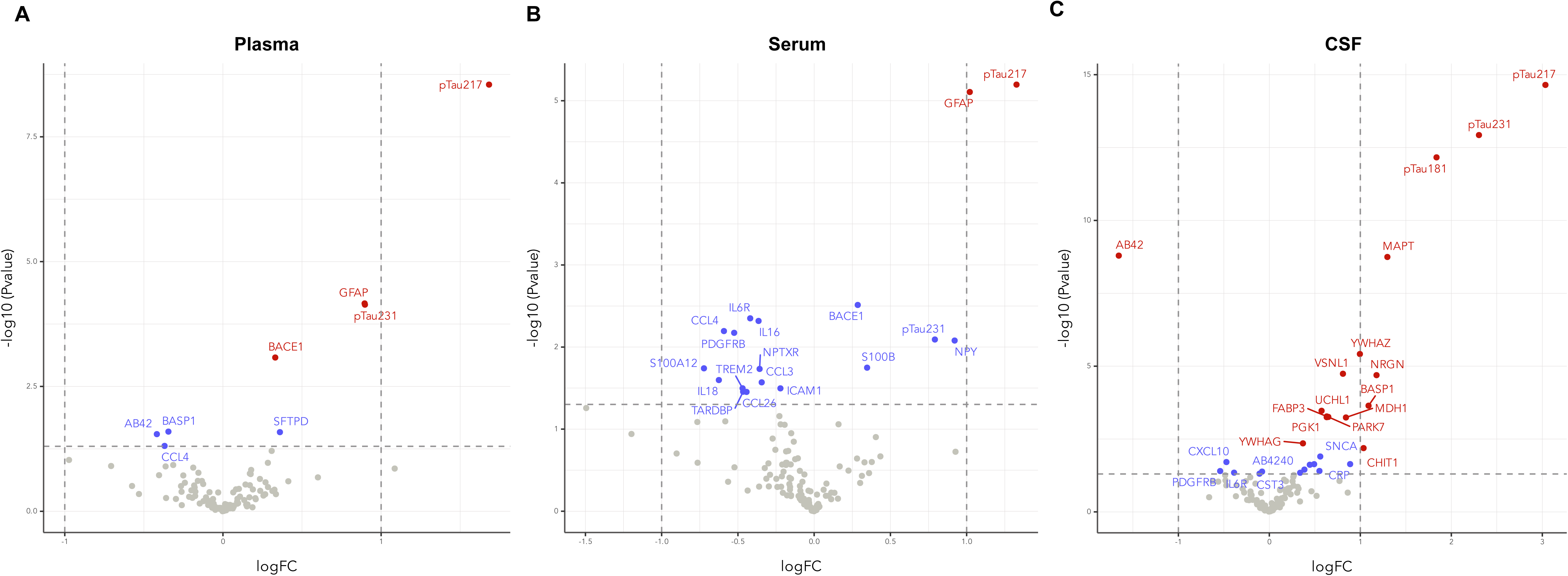
Differential protein changes between biologically determined AD compared to non-AD (cohort 1) in plasma (**A**), serum (**B**) and CSF (**C**). In the volcano plots the red coloured dots indicate proteins significantly different between groups after FDR correction for multiple comparisons, whilst blue coloured dots indicate proteins nominally significant. The horizontal dashed line indicates the unadjusted *P* value threshold of 0.05.

### NULISAseq measurements of p-tau217

In cohort 1, we also compared NULISAseq multiplex p-tau217 measures to other high-performing p-tau217 singleplex immunoassays previously described in the literature (**Suppl. Fig. 7**). We found a high correlation between NULISAseq p-tau217 levels and Janssen p-tau217+ (r = 0.924, *P* < 0.001), ALZpath p-tau217 (r = 0.924, *P* < 0.001), Lumipulse p-tau217 (r = 0.936, *P* < 0.001) and mass spectrometry p-tau217 measures (r = 0.925, *P* < 0.001) with differing fold-changes but similar diagnostic accuracy. NULISAseq and NULISA singleplex measures of p-tau217 strongly correlated (r = 0.906, *P* < 0.00). We observed a larger fold-change for the NULISA singleplex compared to NULISAseq but this did not translate to statistically different diagnostic accuracy.

### NULISAseq in patients with MCI due to AD (A**β**+) compared to MCI unlikely due to AD (A**β**-)

In the plasma of patients with MCI due to AD compared to MCI unlikely due to AD (cohort 2), we observed 17 differentially changed proteins (**Fig. 2A**) but only the upregulation of p-tau217 protein that passed multiple testing correction (p-tau217, logFC = 1.49, *P_adj_* < 0.001); **Suppl. Fig. 8**). Of the nominally significant proteins (**Suppl. Fig. 9**), several cytokines and immune-related proteins (IL6, logFC = -0.668, *P* = 0.001; TNF, logFC = -0.299, *P* = 0.017; CRP, logFC = -0.45, *P =* 0.018; IL15, logFC = -0.23, *P* = 0.027; IL9, logFC = 0.54, *P* = 0.49), amyloid peptides (Aβ38, logFC = -0.45, *P* = 0.004; Aβ42, logFC = -0.43, *P* = 0.011) and proteins implicated in synucleinopathies (PARK7, logFC = -0.912, *P* = 0.027; FABP3, logFC = -0.370, *P* = 0.036) were down-regulated. GFAP (logFC = 0.521, *P* = 0.002) and p-tau231(logFC = 0.611, *P* = 0.004) were up-regulated. In addition, despite not being statistically significant, oligomeric α-synuclein (Oligo-SNCA) had a larger fold-change than p-tau217 (Oligo-SNCA, logFC = -1.54, *P_adj_* > 0.05; **Fig. 2A**). The results for all proteins measured in the multiplex for plasma are displayed in **Suppl. Table 4**.

**Figure 2.**
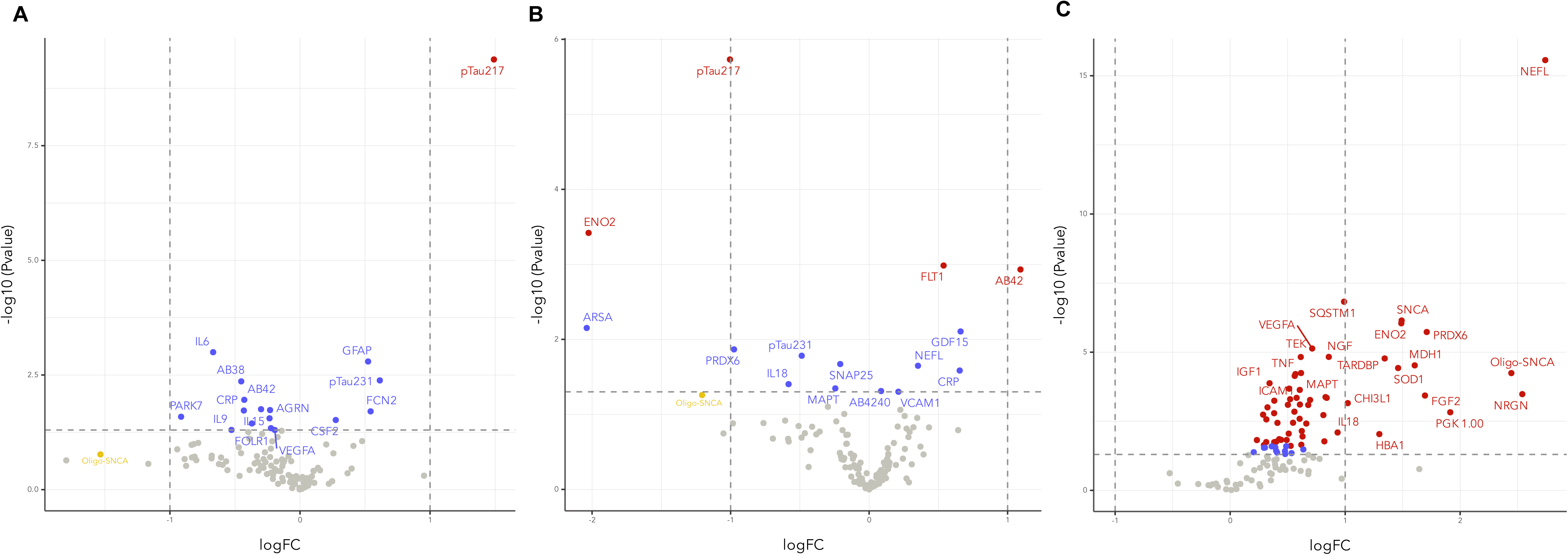
Differential plasma protein changes between MCI+ and MCI- (**A**), LB+ vs AD (**B**) and *GRN*+ and *GRN*- (**C**). In the volcano plots the red coloured dots indicate proteins significantly different between groups after FDR correction for multiple comparisons, whilst blue coloured dots indicate proteins nominally significant. The horizontal dashed line indicates the unadjusted *P* value threshold of 0.05.

### NULISAseq in LB-SAA+ patients compared to AD-SAA-patients

In the plasma of symptomatic patients with and without α-synuclein pathology, determine by CSF SAA, we observed 15 differentially changed proteins (**Fig. 2B**), of which, two proteins upregulated (Aβ42, logFC = 1.09, P*_adj_* = 0.036; vascular endothelial growth factor receptor [FLT1], logFC = 0.537, P*_adj_* = 0.036) and two downregulated (p-tau217, logFC = 1.01, P*_adj_* < 0.001; Neuron specific enolase function [ENO2], logFC = -2.02, P*_adj_* = 0.023; **Suppl. Fig. 10**) passed multiple testing correction. The boxplots of nominally significant proteins are shown in **Suppl. Fig. 11**. We also selectively examined proteins in the CNS panel which were related to α-synuclein (α-synuclein, Oligo-SNCA, phosphorylated α-synuclein-129; **Suppl. Fig. 12**). Interestingly, we found a non-significant change of Oligo-SNCA but with a similar fold-change to p-tau217 (logFC = -1.05, P > 0.05). The results for all proteins measured in the multiplex for plasma are displayed in **Suppl. Table 5**.

### NULISAseq in *GRN* mutation carriers compared to non-carriers

In the plasma of *GRN* mutation carriers we observed 65 differentially upregulated proteins (**Fig. 2C**), of which, 55 proteins passed multiple testing correction compared to non-mutation carriers (**Suppl. Table 6**). The most significantly changed protein was NfL (logFC = 2.74, P*_adj_* < 0.001; **Suppl. Fig. 13**). Notably, Neurogranin (logFC = 2.54, P*_adj_* < 0.001; **Suppl. Fig. 14**), Oligo-SNCA (logFC = 2.46, P*_adj_* < 0.001; **Suppl. Fig. 14**) demonstrated similar fold-changes to NfL. Further, SOD1 (logFC = 2.46, P*_adj_* < 0.001; **Suppl. Fig. 14**) and SQSTM1 (logFC = 2.46, P*_adj_* < 0.001; **Suppl. Fig. 14**) and TDP-43 (logFC = 2.46, P*_adj_* < 0.001; **Suppl. Fig. 14**) all showed significantly different changes between groups.

### Detectability differences between plasma and serum

As alluded to above, commonalities, but many more differences, are observed between plasma and serum when comparing diagnostic groups. To illustrate this further, we correlated the same proteins detected by NULISA^TM^ between plasma and serum (**Suppl. Fig. 15**; **Suppl. Table. 7**). Interestingly, 30.9% of proteins were not significantly correlated between plasma and serum. Of note, p-tau181 (r = 0.019), p-tau231 (r = 0.187), Aβ38 (r = 0.172), Aβ42 (r = 0.101), t-tau (r = 0.161), TDP-43 (r = -0.058), pTDP-43 (r = 0.005), all had poor and non-significant relationships between plasma and serum. Within the significantly correlated proteins, a wide range of correlation coefficients were overserved (r = 0.33-0.971). NfH (r = 0.971; P = 2.4^-25^) and NfL (r = 0.93; P = 4.1^-17^) were among the most highly correlated proteins. P-tau217 (r = 0.662; P = 3.6^-6^) and GFAP (r = 0.887; P = 2.7^-14^) has significant correlations between plasma and serum. Next, we compared the correlations between plasma, serum, and CSF. We observed that 25.6% of proteins were significantly correlated (P < 0.05) between plasma and CSF (**Suppl. Fig. 16**; **Suppl. Table. 8**). The most significantly correlated proteins included CRP (r = -898; P = 5.2^-13^), PDGFRβ (r = 0.728; P = 9.9^-8^), p-tau217 (r = 0.721; P = 1.5^-7^) and NfL (r = 0.701; P = 4.8^-7^). In serum (**Suppl. Fig. 17**; **Suppl. Table. 9**), only 16.7% of proteins were significantly correlated (P < 0.05) between serum and CSF. Like plasma, CRP (r = -887; P = 2.34^-14^), PDGFRβ (r = 0.634; P = 1.1^-6^) and NfL (r = 0.681; P = 1.3^-6^) were amongst the other tightly associated.

## DISCUSSION

The findings from this series of pilot studies highlight the potential of a new generation of multi-protein measurement platforms in enhancing the detection and characterization of biomarkers for neurodegenerative diseases, particularly AD. While mass spectrometry analyses in blood remains to be the only truly unbiased discovery approach, it is hampered by the presence of highly abundant proteins. In contrast, new high plex antibody or aptamer technologies with attomolar sensitivity (e.g., NULISA™) demonstrates a novel approach by combining robust single analytes with known utility (e.g., p-tau217 and NfL) with a multi-analyte screening capacity in low sample volume (< 25 μl).

In AD-related cohorts, this study confirmed the utility of phosphorylated tau (p-tau217) as the leading biomarker in both plasma and serum. The consistent upregulation of p-tau217 across different biological matrices (plasma, serum, and CSF), the correlation within such matrices, and with the NULISA™ p-tau217 measures highly correlated with established single plex immunoassays and mass spectrometry measures, underscores the reliability of the biomarker and this novel multiplex measurement format. In concordance with the previous work, p-tau231 and GFAP were seen as ancillary biomarkers in AD. Fundamentally, the NULISA™ platform also revealed a contingent of biomarkers differentially expressed proteins in AD and MCI which is more representative of the complexity of the disease and the potential for a multi-marker approach to improve underlying pathophysiology of the disease. In participants of biologically determined AD compared to non-AD we measured the multiplex panel in plasma, serum, and CSF. It is generally considered that plasma is the preferred matrix for AD blood biomarkers ^43^, with lower levels of certain key biomarkers serum ^44^, and in the case of Aβ42, weak correlations between the matrices ^44^. Comparing plasma and serum, we reveal differences in significant biomarkers changes between AD and non-AD but also some commonality *e.g.,* p-tau217 and GFAP. Further, we show that approximately one-third of analyzed proteins were not significantly correlated between plasma and serum (correlation coefficient < 0.3) and this illustrates the importance of considering both plasma and serum as complimentary biofluids, not interchangeable biofluids, for a thorough novel biomarker discovery in blood.

The study also explored biomarker profiles in other neurodegenerative conditions in an endophenotype approach, such as Lewy body disease and *GRN* mutation carriers, one of the most common forms of genetic FTD. In LB pathology (SAA+ but Aβ-) compared with AD (SAA-but Aβ+), we confirmed the importance of p-tau217 but also Aβ42 as discriminatory biomarker between AD and non-AD pathologies. However, significant changes in proteins vascular endothelial growth factor receptor, enolase 2 and GDF15 were also observed. A non-significant but noticeable change in oligomeric α-synuclein (Oligo-SNCA) is a promising advancement for LB-related biomarkers in plasma was also seen. Changes in Oligo-SNCA was also observed in MCI-patients compared to MCI+ patients as well as *GRN* mutation carriers. The *GRN* mutation carriers pilot study, observed a remarkable number of upregulation proteins, including NfL, Neurogranin, and TDP-43. The significant fold changes in these proteins indicate their potential utility in identifying and monitoring *GRN*-related pathologies and possible translatable blood biomarkers for sporadic forms of FTD.

Despite these promising results, the study acknowledges several limitations. The group comparisons included in this pilot study are not sufficiently powered to draw definitive conclusions regarding the novel biomarkers identified or the proteins that did not reach statistical significance but exhibited change, which were numerous. This limitation underscores the necessity for larger, more robust cohorts to validate these preliminary findings. Yet, the independent validation of p-tau217, GFAP, and NfL in AD and *GRN* mutations carriers respectively, which were unbiasedly ranked the highest, significantly enhances the credibility of these findings.

In conclusion, this study presents a promising new direction for clinical proteomics that utilises CNS dedicated multiplex panels which incorporate robust known targets but simultaneously allowing for novel discovery. Such advances will offer the blood-based diagnostic and prognostic tools for unmet needs within neurodegenerative disorders.

## Data Availability

All data produced in the present study are available upon reasonable request to the authors

## Sources of Funding

HZ is a Wallenberg Scholar and a Distinguished Professor at the Swedish Research Council supported by grants from the Swedish Research Council (#2023-00356; #2022-01018 and #2019-02397), the European Union’s Horizon Europe research and innovation programme under grant agreement No 101053962, Swedish State Support for Clinical Research (#ALFGBG-71320), the Alzheimer Drug Discovery Foundation (ADDF), USA (#201809-2016862), the AD Strategic Fund and the Alzheimer’s Association (#ADSF-21-831376-C, #ADSF-21-831381-C, #ADSF-21-831377-C, and #ADSF-24-1284328-C), the European Partnership on Metrology, co-financed from the European Union’s Horizon Europe Research and Innovation Programme and by the Participating States (NEuroBioStand, #22HLT07), the Bluefield Project, Cure Alzheimer’s Fund, the Olav Thon Foundation, the Erling-Persson Family Foundation, Familjen Rönströms Stiftelse, Stiftelsen för Gamla Tjänarinnor, Hjärnfonden, Sweden (#FO2022-0270), the European Union’s Horizon 2020 research and innovation programme under the Marie Skłodowska-Curie grant agreement No 860197 (MIRIADE), the European Union Joint Programme – Neurodegenerative Disease Research (JPND2021-00694), the National Institute for Health and Care Research University College London Hospitals Biomedical Research Centre, and the UK Dementia Research Institute at UCL (UKDRI-1003). APi has been supported by grants of Airalzh Foundation AGYR2021 Life-Bio Grant, The LIMPE-DISMOV Foundation Segala Grant 2021, the Italian Ministry of University and Research PRIN COCOON (2017MYJ5TH) and PRIN 2021 RePlast (20202THZAW), the H2020 IMI IDEA-FAST (ID853981), Italian Ministry of Health, Grant/Award Number: RF-2018-12366209 and PNRR-Health PNRR-MAD-2022-12376110. CTo is supported by the Ministry of Health PRIN 2021 RePlast.

APa has been supported by grants of the Italian Ministry of University and Research PRIN COCOON (2017MYJ5TH) and PRIN 2021 RePlast (20202THZAW), the H2020 IMI IDEA-FAST (ID853981), Italian Ministry of Health, Grant/Award Number: RF-2018-12366209 and PNRR-Health PNRR-MAD-2022-12376110.

MS-C receives funding from the European Research Council (ERC) under the European Union’s Horizon 2020 research and innovation programme (Grant agreement No. 948677); ERA PerMed (ERAPERMED2021-184); Project “PI19/00155” and “PI22/00456, funded by Instituto de Salud Carlos III (ISCIII) and co-funded by the European Union; and from a fellowship from “la Caixa” Foundation (ID 100010434) and from the European Union’s Horizon 2020 research and innovation programme under the Marie Skłodowska-Curie grant agreement No 847648 (LCF/BQ/PR21/11840004).

## Disclosures

HZ has served at scientific advisory boards and/or as a consultant for Abbvie, Acumen, Alector, Alzinova, ALZPath, Amylyx, Annexon, Apellis, Artery Therapeutics, AZTherapies, Cognito Therapeutics, CogRx, Denali, Eisai, LabCorp, Merry Life, Nervgen, Novo Nordisk, Optoceutics, Passage Bio, Pinteon Therapeutics, Prothena, Red Abbey Labs, reMYND, Roche, Samumed, Siemens Healthineers, Triplet Therapeutics, and Wave, has given lectures in symposia sponsored by Alzecure, Biogen, Cellectricon, Fujirebio, Lilly, Novo Nordisk, and Roche, and is a co-founder of Brain Biomarker Solutions in Gothenburg AB (BBS), which is a part of the GU Ventures Incubator Program (outside submitted work).

BB has served at scientific advisory boards for Alector, Alexion, AviadoBio, Denali, Lilly/Prevail, UCB and Wave.

APi received consultancy/speaker fees from Abbvie, Bial, Lundbeck, Roche and Zambon pharmaceuticals.

APa received grant support from Ministry of Health (MINSAL) and Ministry of Education, Research and University (MIUR), from CARIPLO Foundation; personal compensation as a consultant/scientific advisory board member for Biogen, Lundbeck, Roche, Nutricia, General Healthcare (GE).

MS-C has given lectures in symposia sponsored by Almirall, Eli Lilly, Novo Nordisk, Roche Diagnostics, and Roche Farma; received consultancy fees (paid to the institution) from Roche Diagnostics; and served on advisory boards of Eli Lilly, Grifols and Roche Diagnostics. He was granted a project and is a site investigator of a clinical trial (funded to the institution) by Roche Diagnostics. In-kind support for research (to the institution) was received from ADx Neurosciences, Alamar Biosciences, Avid Radiopharmaceuticals, Eli Lilly, Fujirebio, Janssen Research & Development, and Roche Diagnostics.

## Notes

### Funding Statement

This study did not receive any funding

### Author Declarations

Ethics committee of Wolfson Centre (Wolfson Centre CSF study 12/0344) gave ethical approval for this work Ethics committee of Institut Hospital del Mar Investigacions Mediques (project code 2014/5638) gave ethical approval for this work Ethics committee of Center for Neurodegenerative Disorders, University of Brescia ((NP 1471 and NP 2189)gave ethical approval for this work

