## Supplemental, methods, tables and figures for "Biomarker Discovery in Alzheimer’s and Neurodegenerative Diseases using Nucleic Acid-Linked Immuno-Sandwich Assay"

^1^ Department of Psychiatry and Neurochemistry, Institute of Neuroscience & Physiology, the Sahlgrenska Academy at the University of Gothenburg, Mölndal, Sweden; ^2^ Banner Alzheimer's Institute and University of Arizona, Phoenix, AZ, USA; ^3^ Banner Sun Health Research Institute, Sun City, AZ 85351, USA; ^4^ Hospital del Mar Research Institute, Barcelona, Spain; ^5^ Barcelonaβeta Brain Research Center (BBRC), Pasqual Maragall Foundation, Barcelona, Spain; ^6^ Servei de Neurologia, Hospital del Mar, Barcelona, Spain;^7^ Department of Medicine and Life Sciencesces, Universitat Pompeu Fabra,  Barcelona, Spain; ^8^ Department of Medicine, Universitat Autònoma de Barcelona, Barcelona, Spain; ^9^ Dementia Research Centre, UCL Queen Square Institute of Neurology, University College London, London, UK; ^10^ UK Dementia Research Institute at UCL, London, UK; ^11^ Department of Neurology, Weill Cornell Medicine and New York - Presbyterian, New York, New York; ^12^ Department of Neurology, Florida Atlantic University, Charles E. Schmidt College of Medicine, Boca Raton, Florida; ^13^ Department of Neurosciences, Biomedicine, and Movement Sciences, Policlinico G. B. Rossi, University of Verona, 37134, Verona, Italy; ^14^ Clinical Investigation in Laboratory, Maggiore Hospital ASST-Crema, Crema, Italy; ^15^ Department of Clinical and Experimental Sciences, Neurology Unit, University of Brescia, Brescia, Italy; ^16^ Department of Continuity of Care and Frailty, Azienda Socio Sanitaria Territoriale (ASST) Spedali Civili, Brescia, Italy; ^17^ Laboratory of Digital Neurology and Biosensors, University of Brescia, Brescia, Italy; ^18^ Brain Health Center, University of Brescia, Brescia, Italy; ^19^ Department of Neuroscience, Biomedicine and Movement Sciences, University of Verona, Verona, Italy; ^20^ Centro de Investigación Biomédica en Red de Fragilidad y Envejecimiento Saludable (CIBERFES), Madrid, Spain; ^21^ Clinical Neurochemistry Laboratory, Sahlgrenska University Hospital, Mölndal, Sweden; ^22^ Paris Brain Institute, ICM, Pitié-Salpêtrière Hospital, Sorbonne University, Paris, France; ^23^ Neurodegenerative Disorder Research Center, Division of Life Sciences and Medicine, and Department of Neurology, Institute on Aging and Brain Disorders, University of Science and Technology of China and First Affiliated Hospital of USTC, Hefei, PR China; ^24^ Hong Kong Center for Neurodegenerative Diseases, Clear Water Bay, Hong Kong, China; ^25^ Wisconsin Alzheimer's Disease Research Center, University of Wisconsin School of Medicine and Public Health, University of Wisconsin-Madison, Madison, WI, USA

**Table of contents**

**Supplementary tables**

**Table S1**- Comparison of plasma protein expression between AD and non-AD (first 50 results by *P* value).

**Table S2**- Comparison of serum protein expression between AD and non-AD (first 50 results by *P* value).

**Table S3**- Comparison of CSF protein expression between AD and non-AD (first 50 results by *P* value).

**Table S4**- Comparison of plasma protein expression between MCI+ and MCI- (first 50 results by *P* value).

**Table S5**- Comparison of plasma protein expression between LB+ and AD (first 50 results by *P* value).

**Table S6**- Comparison of plasma protein expression between *GRN*+ and *GRN*- (first 50 results by *P* value).

**Table S7**- Correlation of the same protein targets in plasma and serum (ranked by *P* value)

**Table S8**- Correlation of the same protein targets in plasma and CSF (ranked by *P* value)

**Table S9**- Correlation of the same protein targets in plasma and serum (ranked by *P* value)

**Supplementary figures**

**Figure S1.** Plasma proteins passing multiple testing correction in biologically determined AD compared to non-AD (cohort 1).

**Figure S2.** Nominally significant plasma proteins in biologically determined AD compared to non-AD (cohort 1).

**Figure S3.** Serum proteins passing multiple testing correction in biologically determined AD compared to non-AD (cohort 1).

**Figure S4.** Nominally significant serum proteins in biologically determined AD compared to non-AD (cohort 1).

**Figure S5.** CSF proteins passing multiple testing correction in biologically determined AD compared to non-AD (cohort 1).

**Figure S6**. Nominally significant CSF proteins in biologically determined AD compared to non-AD (cohort 1).

**Figure S7**. Comparison between NULISAseq plasma pTau217 and other assays.

**Figure S8.** Plasma proteins passing multiple testing correction in MCI Aβ+ compared to MCI Aβ-.

**Figure S9.** Nominally significant plasma proteins in MCI Aβ+ compared to MCI Aβ-.

**Figure S10.** Plasma proteins passing multiple testing correction in LB+ compared to AD.

**Figure S11.** Nominally significant plasma proteins in LB+ compared to AD.

**Figure S12.** Plasma proteins of interest compared between LB+ and AD.

**Figure S13.** Plasma proteins passing multiple testing correction in *GRN*+ compared to *GRN*-.

**Figure S14.** Plasma proteins of interest compared between *GRN*+ and *GRN*-.

**Figure S15** - Correlation of the same protein targets in plasma and serum (ranked Correlation coefficient)

**Figure S16** - Correlation of the same protein targets in plasma and serum (ranked Correlation coefficient)

**Figure S17** - Correlation of the same protein targets in plasma and serum (ranked Correlation coefficient)

**Table S1**- Comparison of plasma protein expression between AD and non-AD in cohort 1 (first 50 results by *P* value).

| ID | UniProtID | logFC | *t* | *B* | *P* value | Adj. *P* value |
| --- | --- | --- | --- | --- | --- | --- |
| pTau217 | P10636 | 1.68243151 | 7.65514351 | 11.1397931 | 2.84E-09 | 3.36E-07 |
| GFAP | P14136 | 0.89576973 | 4.45761813 | 1.28854857 | 6.87E-05 | 0.00285764 |
| pTau231 | P10636 | 0.89777693 | 4.43959288 | 1.22027634 | 7.27E-05 | 0.00285764 |
| BACE1 | P56817 | 0.33070934 | 3.62299917 | -1.0898619 | 0.00083191 | 0.02454131 |
| BASP1 | P80723 | -0.3447705 | -2.3255401 | -4.2620102 | 0.02536532 | 0.48067366 |
| SFTPD | P35247 | 0.36021297 | 2.31472682 | -4.2823952 | 0.02601139 | 0.48067366 |
| AB42 | P05067 | -0.4179194 | -2.2749878 | -4.3340006 | 0.02851454 | 0.48067366 |
| CCL4 | P13236 | -0.3689988 | -2.0317656 | -4.8106376 | 0.04906712 | 0.72374009 |
| POSTN | Q15063 | 0.30808589 | 1.92309067 | -5.0389012 | 0.06183166 | 0.81068172 |
| MME | P08473 | -0.97324 | -1.7177449 | -5.395637 | 0.09381732 | 0.9966394 |
| pTau181 | P10636 | 0.28406777 | 1.65009493 | -5.4909659 | 0.1069987 | 0.9966394 |
| PDGFRB | P09619 | -0.314521 | -1.5969889 | -5.5659395 | 0.11838344 | 0.9966394 |
| CRP | P02741 | -0.7070814 | -1.5727272 | -5.6242237 | 0.12390371 | 0.9966394 |
| TREM2 | Q9NZC2 | -0.3625838 | -1.5623343 | -5.6329837 | 0.12633144 | 0.9966394 |
| IL6R | P08887 | -0.1878761 | -1.532207 | -5.6745903 | 0.13356794 | 0.9966394 |
| NEFH | P12036 | 1.08588827 | 1.50880157 | -5.7187454 | 0.1394519 | 0.9966394 |
| PGF | P49763 | 0.17847253 | 1.4742682 | -5.7773369 | 0.14846031 | 0.9966394 |
| TNF | P01375 | -0.1550728 | -1.40046 | -5.8432747 | 0.16930526 | 0.9966394 |
| SNAP25 | P60880 | 0.17025212 | 1.37955187 | -5.8890562 | 0.17560724 | 0.9966394 |
| PTN | P21246 | 0.1753795 | 1.36611722 | -5.9014702 | 0.17975159 | 0.9966394 |
| IL16 | Q14005 | -0.2507351 | -1.3296976 | -5.9492979 | 0.19138434 | 0.9966394 |
| SQSTM1 | Q13501 | -0.4039895 | -1.3361665 | -5.9530043 | 0.18928048 | 0.9966394 |
| PDLIM5 | Q96HC4 | 0.60047048 | 1.27653618 | -6.0066181 | 0.20935544 | 0.9966394 |
| S100B | P04271 | 0.23386383 | 1.23244941 | -6.0769993 | 0.22519891 | 0.9966394 |
| FABP3 | P05413 | -0.1587092 | -1.1782927 | -6.1240493 | 0.24584241 | 0.9966394 |
| AGRN | O00468 | -0.1330203 | -1.1748813 | -6.1292233 | 0.247187 | 0.9966394 |
| CD40LG | P29965 | 0.4134112 | 1.16628366 | -6.1490518 | 0.25062017 | 0.9966394 |
| SFRP1 | Q8N474 | 0.2690654 | 1.13640349 | -6.1939293 | 0.2627595 | 0.9966394 |
| REST | Q13127 | -0.2407614 | -1.12284 | -6.1978163 | 0.268407 | 0.9966394 |
| NPY | P01303 | 0.14256686 | 1.070425 | -6.2258875 | 0.29102774 | 0.9966394 |
| ICAM1 | P05362 | -0.1249944 | -1.0862994 | -6.252303 | 0.28403515 | 0.9966394 |
| CCL3 | P10147 | -0.1820432 | -1.0419187 | -6.2733379 | 0.30389962 | 0.9966394 |
| VSNL1 | P62760 | 0.17026804 | 1.04901982 | -6.2907177 | 0.30066089 | 0.9966394 |
| IL5 | P05113 | -0.2649183 | -1.023311 | -6.2997963 | 0.31250022 | 0.9966394 |
| PARK7 | Q99497 | -0.5747687 | -1.0257999 | -6.3127942 | 0.31134025 | 0.9966394 |
| NRGN | Q92686 | 0.28468994 | 0.99212968 | -6.3533962 | 0.32728278 | 0.9966394 |
| CCL11 | P51671 | 0.10093057 | 0.93520249 | -6.3642812 | 0.35545592 | 0.9966394 |
| SLIT2 | O94813 | 0.08251242 | 0.91919466 | -6.3659981 | 0.36366103 | 0.9966394 |
| FOLR1 | P15328 | -0.1789906 | -0.9791799 | -6.3675004 | 0.33355858 | 0.9966394 |
| MAPT | P10636 | 0.18919334 | 0.91598973 | -6.41209 | 0.36533173 | 0.9966394 |
| IFNG | P01579 | 0.32568519 | 0.90802335 | -6.4296075 | 0.3694726 | 0.9966394 |
| CD63 | P08962 | -0.1995834 | -0.8861406 | -6.4507284 | 0.38100239 | 0.9966394 |
| CCL2 | P13500 | -0.1743654 | -0.842569 | -6.4518199 | 0.40463462 | 0.9966394 |
| S100A12 | P80511 | -0.2617003 | -0.8388548 | -6.4617291 | 0.40669045 | 0.9966394 |
| VEGFD | O43915 | 0.06506711 | 0.8190074 | -6.4626141 | 0.41777376 | 0.9966394 |
| CRH | P06850 | 0.31274531 | 0.86616937 | -6.4642278 | 0.39172312 | 0.9966394 |
| CXCL8 | P10145 | 0.23579937 | 0.87022533 | -6.4709858 | 0.38953058 | 0.9966394 |
| AB4240 | P123 | -0.0301305 | -0.8150959 | -6.4851249 | 0.4199821 | 0.9966394 |
| TEK | Q02763 | -0.0848925 | -0.8255047 | -6.4935444 | 0.41412131 | 0.9966394 |
| NPTXR | O95502 | -0.1070721 | -0.6392729 | -6.5235252 | 0.52640264 | 0.9966394 |

**Table S2**- Comparison of serum protein expression between AD and non-AD in cohort 1 (first 50 results by *P* value).

| ID | UniProtID | logFC | *t* | *B* | *P* value | Adj. *P* value |
| --- | --- | --- | --- | --- | --- | --- |
| pTau217 | P10636 | 1.32674659 | 5.19653673 | 3.79389929 | 6.36E-06 | 0.00048141 |
| GFAP | P14136 | 1.02026848 | 5.13163339 | 3.60170785 | 7.83E-06 | 0.00048141 |
| BACE1 | P56817 | 0.28574015 | 3.15170457 | -1.9878357 | 0.00307118 | 0.11389169 |
| IL6R | P08887 | -0.4195099 | -3.0131205 | -2.3417834 | 0.0044693 | 0.11389169 |
| IL16 | Q14005 | -0.364633 | -2.9857114 | -2.3895248 | 0.00480891 | 0.11389169 |
| CCL4 | P13236 | -0.5916711 | -2.8787184 | -2.6619046 | 0.00638897 | 0.11389169 |
| PDGFRB | P09619 | -0.5246538 | -2.8600409 | -2.6997975 | 0.00670847 | 0.11389169 |
| pTau231 | P10636 | 0.79148932 | 2.78766205 | -2.8627821 | 0.00809263 | 0.11389169 |
| NPY | P01303 | 0.921005 | 2.77625712 | -2.8902198 | 0.00833354 | 0.11389169 |
| S100B | P04271 | 0.34699461 | 2.47081186 | -3.5910049 | 0.01783674 | 0.18934934 |
| S100A12 | P80511 | -0.7226721 | -2.463605 | -3.5937374 | 0.0181652 | 0.18934934 |
| NPTXR | O95502 | -0.3577982 | -2.4562854 | -3.5943815 | 0.01847311 | 0.18934934 |
| IL18 | Q14116 | -0.6250294 | -2.3252012 | -3.8899468 | 0.02523404 | 0.23642842 |
| CCL3 | P10147 | -0.3440906 | -2.2972943 | -3.8986543 | 0.02691055 | 0.23642842 |
| TREM2 | Q9NZC2 | -0.4694577 | -2.2248034 | -4.0827687 | 0.03181648 | 0.24058007 |
| ICAM1 | P05362 | -0.2213658 | -2.22401 | -4.0877187 | 0.03185468 | 0.24058007 |
| CCL26 | Q9Y258 | -0.4434529 | -2.1801455 | -4.14573 | 0.03520684 | 0.24058007 |
| TARDBP | Q13148 | -0.4659201 | -2.1826069 | -4.1663601 | 0.03501195 | 0.24058007 |
| PGK1 | P00558 | -1.4947583 | -1.9740672 | -4.5730382 | 0.0553273 | 0.35817146 |
| TNF | P01375 | -0.2278038 | -1.8658402 | -4.7242254 | 0.06940496 | 0.42684049 |
| ANXA5 | P08758 | -0.5828074 | -1.7943104 | -4.8549455 | 0.08034534 | 0.43534379 |
| CRP | P02741 | -0.7661674 | -1.785205 | -4.8983361 | 0.08183365 | 0.43534379 |
| SNAP25 | P60880 | 0.15965326 | 1.75311494 | -4.9301897 | 0.08723968 | 0.43534379 |
| FOLR1 | P15328 | -0.2203494 | -1.752323 | -4.9304537 | 0.08737745 | 0.43534379 |
| FLT1 | P17948 | -0.1936706 | -1.7459968 | -4.9530815 | 0.08848451 | 0.43534379 |
| AGRN | O00468 | -0.1505404 | -1.62458 | -5.1160214 | 0.1121029 | 0.52157158 |
| PARK7 | Q99497 | -1.1980283 | -1.6136331 | -5.1686457 | 0.11449132 | 0.52157158 |
| pTau181 | P10636 | 0.40431613 | 1.56703207 | -5.2066095 | 0.12500687 | 0.54913733 |
| MDH1 | P40925 | -0.2726312 | -1.5448869 | -5.2409569 | 0.13027112 | 0.55252922 |
| IGFBP7 | Q16270 | -0.1338146 | -1.4975087 | -5.3018649 | 0.14210705 | 0.58263892 |
| pTDP43 | Q13148 | -0.1701598 | -1.3515712 | -5.494824 | 0.18410286 | 0.68192639 |
| TREM1 | Q9NP99 | -0.2544852 | -1.3697211 | -5.5090145 | 0.17843569 | 0.68192639 |
| IL33 | O95760 | -0.1841315 | -1.3433091 | -5.5126081 | 0.18673986 | 0.68192639 |
| NEFH | P12036 | 0.92680886 | 1.3379212 | -5.5419284 | 0.18849998 | 0.68192639 |
| GDNF | P39905 | -0.9020513 | -1.3089697 | -5.5559348 | 0.1980356 | 0.69595367 |
| IFNG | P01579 | 0.43415611 | 1.26076883 | -5.6426443 | 0.2147169 | 0.6994581 |
| TEK | Q02763 | -0.1068355 | -1.2282692 | -5.6842049 | 0.22652036 | 0.6994581 |
| CCL11 | P51671 | 0.28095512 | 1.22447569 | -5.6875941 | 0.22795537 | 0.6994581 |
| NPTX2 | P47972 | -0.1246943 | -1.2127971 | -5.6888687 | 0.23231777 | 0.6994581 |
| REST | Q13127 | -0.1802953 | -1.1510215 | -5.702273 | 0.25655641 | 0.6994581 |
| IL17A | Q16552 | 0.37773415 | 1.16420408 | -5.7476082 | 0.25125738 | 0.6994581 |
| CSF2 | P04141 | -0.1847611 | -1.1505685 | -5.7504953 | 0.25675481 | 0.6994581 |
| CRH | P06850 | 0.33000947 | 1.15823198 | -5.7584048 | 0.2536571 | 0.6994581 |
| MME | P08473 | -0.7643801 | -1.1509714 | -5.7605019 | 0.25659679 | 0.6994581 |
| IL5 | P05113 | -0.2999525 | -1.1401226 | -5.771425 | 0.26103494 | 0.6994581 |
| CCL13 | Q99616 | -0.1849153 | -1.1116046 | -5.7834155 | 0.27295926 | 0.6994581 |
| FABP3 | P05413 | -0.1537871 | -1.1204858 | -5.7891593 | 0.26918574 | 0.6994581 |
| NPTX1 | Q15818 | 0.16739421 | 1.0847639 | -5.8006696 | 0.28452097 | 0.7142057 |
| FCN2 | Q15485 | -0.2096082 | -1.1190463 | -5.8141224 | 0.26981397 | 0.6994581 |
| YWHAZ | P63104 | -0.5228127 | -1.0688333 | -5.8573383 | 0.2915704 | 0.71726317 |

**Table S3**- Comparison of CSF protein expression between AD and non-AD in cohort 1 (first 50 results by *P* value).

| ID | UniProtID | logFC | *t* | *B* | *P* value | Adj. *P* value |
| --- | --- | --- | --- | --- | --- | --- |
| pTau217 | P10636 | 3.03435342 | 12.3977141 | 24.885293 | 2.24E-15 | 2.65E-13 |
| pTau231 | P10636 | 2.30350496 | 10.9288587 | 20.9288617 | 1.18E-13 | 6.95E-12 |
| pTau181 | P10636 | 1.83724058 | 10.3067148 | 19.1744638 | 6.86E-13 | 2.70E-11 |
| AB42 | P05067 | -1.6550807 | -7.7443077 | 11.4533453 | 1.61E-09 | 4.24E-08 |
| MAPT | P10636 | 1.2971381 | 7.71064743 | 11.3179016 | 1.79E-09 | 4.24E-08 |
| YWHAZ | P63104 | 0.99503472 | 5.34138503 | 3.69957368 | 3.83E-06 | 7.53E-05 |
| VSNL1 | P62760 | 0.808747 | 4.85582595 | 2.16998707 | 1.82E-05 | 0.00030023 |
| NRGN | Q92686 | 1.17781584 | 4.82110458 | 2.04641871 | 2.04E-05 | 0.00030023 |
| BASP1 | P80723 | 1.09029356 | 4.04684486 | -0.291277 | 0.00022721 | 0.00297902 |
| UCHL1 | P09936 | 0.57396341 | 3.91323201 | -0.6881887 | 0.00034008 | 0.00401293 |
| FABP3 | P05413 | 0.63090538 | 3.76334089 | -1.1092759 | 0.00053156 | 0.00485892 |
| PARK7 | Q99497 | 0.64804837 | 3.75051727 | -1.1320253 | 0.0005521 | 0.00485892 |
| PGK1 | P00558 | 0.6302407 | 3.74412603 | -1.1385224 | 0.00056262 | 0.00485892 |
| MDH1 | P40925 | 0.84165111 | 3.73587727 | -1.1943061 | 0.00057648 | 0.00485892 |
| YWHAG | P61981 | 0.3691298 | 3.01217229 | -3.1317874 | 0.00445331 | 0.03503269 |
| CHIT1 | Q13231 | 1.03557775 | 2.87084031 | -3.4383185 | 0.00647889 | 0.04778179 |
| SNCA | P37840 | 0.55942304 | 2.61155645 | -4.0977552 | 0.01257858 | 0.08731017 |
| CXCL10 | P02778 | -0.4721216 | -2.4324539 | -4.4822481 | 0.01950118 | 0.12784109 |
| TIMP3 | P35625 | 0.49211591 | 2.36201859 | -4.6177237 | 0.02306258 | 0.1358092 |
| CRP | P02741 | 0.88787209 | 2.36424195 | -4.6492648 | 0.02294175 | 0.1358092 |
| GOT1 | P17174 | 0.44454098 | 2.34212614 | -4.6903525 | 0.02416943 | 0.1358092 |
| SOD1 | P00441 | 0.3853024 | 2.1770923 | -5.0240625 | 0.03535228 | 0.1896168 |
| PDGFRB | P09619 | -0.5408532 | -2.1253699 | -5.1158573 | 0.03969783 | 0.19518102 |
| ENO2 | P09104 | 0.55052654 | 2.12769849 | -5.1348075 | 0.03949252 | 0.19518102 |
| AB4240 | P123 | -0.0800184 | -2.105217 | -5.1622907 | 0.04149016 | 0.19583357 |
| IL6R | P08887 | -0.3876549 | -2.0668403 | -5.232066 | 0.04517625 | 0.1986223 |
| BACE1 | P56817 | 0.33846505 | 2.06410754 | -5.2403252 | 0.04544748 | 0.1986223 |
| CST3 | P01034 | -0.1081603 | -2.0306521 | -5.2982219 | 0.04885947 | 0.20590775 |
| CALB2 | P22676 | 0.26912895 | 1.98093114 | -5.3369736 | 0.05441119 | 0.21387404 |
| NEFH | P12036 | -0.4858045 | -1.9757349 | -5.3863798 | 0.05501881 | 0.21387404 |
| SLIT2 | O94813 | 0.26871864 | 1.96588054 | -5.4251861 | 0.05618725 | 0.21387404 |
| PRDX6 | P30041 | 0.26321344 | 1.90943435 | -5.5296897 | 0.06330101 | 0.2323845 |
| RUVBL2 | Q9Y230 | -0.2098911 | -1.8967743 | -5.5514946 | 0.06498889 | 0.2323845 |
| IL5 | P05113 | 0.16193799 | 1.80510129 | -5.6790883 | 0.07846164 | 0.26300392 |
| IFNG | P01579 | -0.482184 | -1.7774806 | -5.7641015 | 0.08299221 | 0.26300392 |
| CD63 | P08962 | 0.18301776 | 1.74754827 | -5.7708887 | 0.08808531 | 0.26300392 |
| SQSTM1 | Q13501 | 0.25517284 | 1.74165137 | -5.7880363 | 0.08915387 | 0.26300392 |
| IL6 | P05231 | -0.2592682 | -1.7465113 | -5.7921442 | 0.08829631 | 0.26300392 |
| SFRP1 | Q8N474 | 0.77081017 | 1.75491083 | -5.7966306 | 0.08683043 | 0.26300392 |
| SAA1 | P0DJI8 | -0.6217746 | -1.7435629 | -5.8232539 | 0.08881575 | 0.26300392 |
| IL10 | P22301 | -0.5620414 | -1.7284921 | -5.8418868 | 0.09151089 | 0.2633728 |
| TARDBP | Q13148 | -0.1532707 | -1.6463422 | -5.9639973 | 0.10739597 | 0.30173154 |
| CD40LG | P29965 | -0.4740962 | -1.5739595 | -6.0426739 | 0.12326997 | 0.33015738 |
| IL4 | P05112 | -0.3615786 | -1.5815235 | -6.0566577 | 0.12153024 | 0.33015738 |
| CHI3L1 | P36222 | 0.26611784 | 1.56265543 | -6.1053208 | 0.12590748 | 0.33015738 |
| NEFL | P07196 | 0.31198139 | 1.51139369 | -6.133557 | 0.13844504 | 0.35514162 |
| CCL3 | P10147 | 0.22351322 | 1.45574669 | -6.2381489 | 0.153164 | 0.37816547 |
| PSEN1 | P49768 | 0.20668048 | 1.4469901 | -6.2615165 | 0.15558877 | 0.37816547 |
| IL7 | P13232 | 0.32075477 | 1.43173785 | -6.2940825 | 0.15988431 | 0.37816547 |
| PGF | P49763 | -0.2936371 | -1.4304907 | -6.3050761 | 0.16023961 | 0.37816547 |

**Table S4**- Comparison of plasma protein expression between MCI+ and MCI- (first 50 results by *P* value).

| ID | UniProtID | logFC | *t* | *B* | *P v*alue | Adj. *P* value |
| --- | --- | --- | --- | --- | --- | --- |
| pTau217 | P10636 | 1.49084953 | 8.24616777 | 12.9955608 | 4.17E-10 | 4.92E-08 |
| IL6 | P05231 | -0.6678983 | -3.5536975 | -1.3321434 | 0.00100563 | 0.05933243 |
| GFAP | P14136 | 0.52342787 | 3.38975674 | -1.8020134 | 0.00160291 | 0.06304763 |
| pTau231 | P10636 | 0.61359182 | 3.04425922 | -2.7026424 | 0.00414636 | 0.10265746 |
| AB38 | P05067 | -0.4520076 | -3.0263695 | -2.7580663 | 0.00434989 | 0.10265746 |
| AB42 | P05067 | -0.4286731 | -2.6694332 | -3.6135894 | 0.01098583 | 0.21605459 |
| TNF | P01375 | -0.2995502 | -2.4779621 | -4.0332394 | 0.01760959 | 0.23126887 |
| AGRN | O00468 | -0.2303654 | -2.4603674 | -4.0723138 | 0.01837253 | 0.23126887 |
| CRP | P02741 | -0.4318147 | -2.4508565 | -4.068974 | 0.01879733 | 0.23126887 |
| FCN2 | Q15485 | 0.54274533 | 2.4334265 | -4.0962155 | 0.01959906 | 0.23126887 |
| PARK7 | Q99497 | -0.9142411 | -2.3186486 | -4.3672355 | 0.02570627 | 0.27343754 |
| IL15 | P40933 | -0.2344156 | -2.2846528 | -4.4389861 | 0.02780721 | 0.27343754 |
| CSF2 | P04141 | 0.27490441 | 2.24755459 | -4.4375113 | 0.03028399 | 0.27488547 |
| FABP3 | P05413 | -0.3703325 | -2.1719325 | -4.6360105 | 0.03594988 | 0.30300614 |
| FOLR1 | P15328 | -0.2234717 | -2.064581 | -4.8522589 | 0.04559938 | 0.34050389 |
| IL9 | P15248 | -0.5269676 | -2.0236433 | -4.9364632 | 0.04983745 | 0.34050389 |
| VEGFA | P15692 | -0.1936505 | -2.0221626 | -4.9539012 | 0.04999697 | 0.34050389 |
| IGFBP7 | Q16270 | -0.1413628 | -2.0044375 | -4.9452843 | 0.05194127 | 0.34050389 |
| CCL2 | P13500 | -0.3978338 | -1.9740367 | -5.0459772 | 0.05542907 | 0.34424372 |
| IL33 | O95760 | -0.2910771 | -1.9315346 | -5.1056667 | 0.06064387 | 0.35779883 |
| CX3CL1 | P78423 | -0.1719156 | -1.8547655 | -5.2196178 | 0.07113601 | 0.39971663 |
| CCL4 | P13236 | -0.3907628 | -1.8153864 | -5.2977124 | 0.07709329 | 0.41350035 |
| SFRP1 | Q8N474 | 0.47787233 | 1.75327631 | -5.425684 | 0.08734458 | 0.43218854 |
| AB40 | P05067 | -0.2135349 | -1.7500739 | -5.4130267 | 0.08790275 | 0.43218854 |
| SAA1 | P0DJI8 | -0.7826509 | -1.7081421 | -5.5068087 | 0.0955022 | 0.44489002 |
| CXCL1 | P09341 | -0.8158079 | -1.6783764 | -5.53413 | 0.10121313 | 0.44489002 |
| VSNL1 | P62760 | 0.2017937 | 1.66764754 | -5.5548612 | 0.10332833 | 0.44489002 |
| IL18 | Q14116 | -0.8335711 | -1.6566034 | -5.5733952 | 0.10556712 | 0.44489002 |
| IL4 | P05112 | 0.4106815 | 1.6302057 | -5.5899864 | 0.11104025 | 0.45181894 |
| SFTPD | P35247 | -0.4708241 | -1.5932347 | -5.6798736 | 0.11911198 | 0.46850711 |
| CCL3 | P10147 | -0.2240979 | -1.557076 | -5.7147188 | 0.127462 | 0.48517794 |
| CD40LG | P29965 | -0.9439085 | -1.5386816 | -5.7516036 | 0.13189924 | 0.48637845 |
| KLK6 | Q92876 | -0.2698707 | -1.4571412 | -5.8718874 | 0.15301417 | 0.52724823 |
| SQSTM1 | Q13501 | -0.4533676 | -1.4321455 | -5.9269951 | 0.16000075 | 0.52724823 |
| NPTX1 | Q15818 | 0.25676751 | 1.42811868 | -5.9058745 | 0.16114929 | 0.52724823 |
| Oligo-SNCA | P37840 | -1.5345273 | -1.3964907 | -5.9733777 | 0.17040807 | 0.52724823 |
| SNCA | P37840 | -0.678714 | -1.3614023 | -6.0210992 | 0.18114484 | 0.52724823 |
| CXCL10 | P02778 | -0.3312861 | -1.3462096 | -6.0300861 | 0.18594152 | 0.52724823 |
| IGF1 | P05019 | -0.088904 | -1.34282 | -6.0106044 | 0.18702741 | 0.52724823 |
| TARDBP | Q13148 | -0.8103686 | -1.3326412 | -6.0469969 | 0.19032843 | 0.52724823 |
| IL16 | Q14005 | -0.2297843 | -1.3211567 | -6.0305779 | 0.19408257 | 0.52724823 |
| TIMP3 | P35625 | -0.8308845 | -1.3078341 | -6.0889362 | 0.19853186 | 0.52724823 |
| PSEN1 | P49768 | -0.6069135 | -1.3069576 | -6.0915412 | 0.19882654 | 0.52724823 |
| SOD1 | P00441 | -0.6800011 | -1.2812616 | -6.1269418 | 0.20761399 | 0.52724823 |
| REST | Q13127 | -0.280162 | -1.259675 | -6.1231066 | 0.21520909 | 0.52724823 |
| NRGN | Q92686 | -0.887243 | -1.2592835 | -6.1338483 | 0.21535951 | 0.52724823 |
| CCL11 | P51671 | -0.1382335 | -1.2497673 | -6.1349548 | 0.21876899 | 0.52724823 |
| FGF2 | P09038 | -0.7974367 | -1.2259427 | -6.1618929 | 0.22751911 | 0.52724823 |
| ANXA5 | P08758 | -1.7970464 | -1.2200398 | -6.2006368 | 0.2297239 | 0.52724823 |
| VCAM1 | P19320 | -0.1207762 | -1.2108478 | -6.1494776 | 0.23317831 | 0.52724823 |

**Table S5**- Comparison of plasma protein expression between LB+ and AD (first 50 results by *P* value).

| ID | UniProtID | logFC | *t* | *B* | *P* value | Adj. *P* value |
| --- | --- | --- | --- | --- | --- | --- |
| pTau217 | P10636 | -1.0047838 | -5.3469519 | 4.88240041 | 1.85E-06 | 0.00023 |
| ENO2 | P09104 | -2.0253943 | -3.7909443 | -0.1604672 | 0.00038025 | 0.02357531 |
| FLT1 | P17948 | 0.53761162 | 3.46830486 | -1.0716818 | 0.00103553 | 0.03627919 |
| AB42 | P05067 | 1.09261336 | 3.42818312 | -1.1956855 | 0.0011703 | 0.03627919 |
| ARSA | P15289 | -2.038832 | -2.8016165 | -2.8436017 | 0.00704447 | 0.16198751 |
| GDF15 | Q99988 | 0.66022631 | 2.76190445 | -2.9181001 | 0.00783811 | 0.16198751 |
| PRDX6 | P30041 | -0.9753834 | -2.5514974 | -3.422475 | 0.01359141 | 0.24076204 |
| pTau231 | P10636 | -0.4866509 | -2.4744796 | -3.5781179 | 0.01651641 | 0.25600435 |
| SNAP25 | P60880 | -0.2086949 | -2.3712431 | -3.7943353 | 0.02132086 | 0.27856811 |
| NEFL | P07196 | 0.35265903 | 2.34979164 | -3.8268429 | 0.02246517 | 0.27856811 |
| CRP | P02741 | 0.6537743 | 2.28922609 | -3.9849509 | 0.02600318 | 0.29312679 |
| IL18 | Q14116 | -0.5815769 | -2.1099585 | -4.3326573 | 0.03951239 | 0.40829467 |
| MAPT | P10636 | -0.2441972 | -2.0522002 | -4.4256978 | 0.04500542 | 0.41227096 |
| AB4240 | P123 | 0.08616613 | 2.01553027 | -4.5036612 | 0.04883152 | 0.41227096 |
| VCAM1 | P19320 | 0.21132311 | 2.00599295 | -4.5340051 | 0.04987149 | 0.41227096 |
| YWHAZ | P63104 | -1.2059419 | -1.959889 | -4.6416395 | 0.05517918 | 0.42763866 |
| pTau181 | P10636 | -0.2975587 | -1.7896198 | -4.9043227 | 0.07911666 | 0.57708626 |
| NGF | P01138 | 0.2251283 | 1.74388812 | -4.9978235 | 0.08686262 | 0.58217261 |
| BACE1 | P56817 | -0.179414 | -1.6800679 | -5.0885494 | 0.09871447 | 0.58217261 |
| SQSTM1 | Q13501 | 0.25623221 | 1.64891444 | -5.1286522 | 0.10496491 | 0.58217261 |
| SMOC1 | Q9H4F8 | 0.34704854 | 1.61786366 | -5.1791616 | 0.11152043 | 0.58217261 |
| GFAP | P14136 | -0.2371986 | -1.5629335 | -5.2414895 | 0.12390357 | 0.58217261 |
| CNTN2 | Q02246 | -0.6065268 | -1.5756608 | -5.2672476 | 0.12094639 | 0.58217261 |
| HBA1 | P69905 | -0.9833924 | -1.5310707 | -5.3106549 | 0.13159103 | 0.58217261 |
| PARK7 | Q99497 | -0.7625749 | -1.5358296 | -5.3279214 | 0.13042054 | 0.58217261 |
| VGF | O15240 | 0.43226121 | 1.46328724 | -5.3881867 | 0.1491869 | 0.58217261 |
| VEGFD | O43915 | -0.5093568 | -1.4863585 | -5.396016 | 0.14300127 | 0.58217261 |
| VEGFA | P15692 | 0.20600819 | 1.4777827 | -5.4005203 | 0.14526848 | 0.58217261 |
| SNCA | P37840 | -0.4116657 | -1.474916 | -5.4057004 | 0.14604335 | 0.58217261 |
| IL6 | P05231 | 0.31987947 | 1.46539557 | -5.4156621 | 0.14861306 | 0.58217261 |
| FABP3 | P05413 | 0.27040963 | 1.44332296 | -5.4507841 | 0.15470724 | 0.58217261 |
| TEK | Q02763 | -0.1919013 | -1.416749 | -5.4744597 | 0.1622932 | 0.58217261 |
| MME | P08473 | 0.64262 | 1.41740039 | -5.4757338 | 0.16211156 | 0.58217261 |
| CCL26 | Q9Y258 | -0.3571589 | -1.3920341 | -5.4989639 | 0.16962027 | 0.58217261 |
| IL33 | O95760 | 0.29206435 | 1.38665844 | -5.5371333 | 0.17124544 | 0.58217261 |
| ACHE | P22303 | -0.1645201 | -1.3339963 | -5.5373583 | 0.18779762 | 0.58217261 |
| OligoSNCA | P37840 | -1.0515005 | -1.3599916 | -5.560087 | 0.17948497 | 0.58217261 |
| MDH1 | P40925 | -0.4640556 | -1.3641328 | -5.5611348 | 0.17818591 | 0.58217261 |
| BASP1 | P80723 | -0.3815424 | -1.3438467 | -5.5626606 | 0.1846187 | 0.58217261 |
| VSNL1 | P62760 | -0.1542101 | -1.3375968 | -5.5760166 | 0.18662803 | 0.58217261 |
| AB40 | P05067 | 0.28378455 | 1.29690072 | -5.6362433 | 0.20018083 | 0.59727366 |
| PDLIM5 | Q96HC4 | -0.4878668 | -1.2856508 | -5.6596537 | 0.20405237 | 0.59727366 |
| PGK1 | P00558 | -0.6948746 | -1.2679032 | -5.6754541 | 0.21027347 | 0.59727366 |
| IL1B | P01584 | -0.5850009 | -1.2632272 | -5.6947103 | 0.21193581 | 0.59727366 |
| FCN2 | Q15485 | -0.1648655 | -1.1704628 | -5.7405484 | 0.24694852 | 0.63795034 |
| S100A12 | P80511 | -0.5804367 | -1.2146584 | -5.7452877 | 0.22978248 | 0.63317839 |
| KLK6 | Q92876 | 0.17853297 | 1.18727299 | -5.7509562 | 0.24031058 | 0.63795034 |
| CCL11 | P51671 | -0.1615475 | -1.1742101 | -5.7947862 | 0.24545749 | 0.63795034 |
| FGF2 | P09038 | 0.37052548 | 1.11718933 | -5.8336708 | 0.26886062 | 0.68038199 |
| CX3CL1 | P78423 | 0.13417834 | 1.08706529 | -5.8779875 | 0.28183094 | 0.69894074 |

**Table S6**- Comparison of CSF protein expression between *GRN*+ and *GRN*- (first 50 results by *P* value).

| ID | UniProtID | logFC | *t* | *B* | *P* value | Adj. *P* value |
| --- | --- | --- | --- | --- | --- | --- |
| NEFL | P07196 | 2.73999573 | 13.33713 | 26.9812752 | 2.71E-16 | 3.20E-14 |
| SQSTM1 | Q13501 | 0.99014751 | 6.35904918 | 6.83486723 | 1.49E-07 | 8.80E-06 |
| SNCA | P37840 | 1.4912041 | 5.87449166 | 5.30670677 | 7.15E-07 | 2.66E-05 |
| ENO2 | P09104 | 1.48817792 | 5.80322845 | 5.08370853 | 9.00E-07 | 2.66E-05 |
| PRDX6 | P30041 | 1.70946599 | 5.57849309 | 4.35816168 | 1.86E-06 | 4.39E-05 |
| VEGFA | P15692 | 0.71326848 | 5.15006491 | 2.96460501 | 7.35E-06 | 0.00014453 |
| NGF | P01138 | 0.85770198 | 4.93126155 | 2.26937617 | 1.48E-05 | 0.0002204 |
| TEK | Q02763 | 0.61394465 | 4.92737403 | 2.26687448 | 1.49E-05 | 0.0002204 |
| TARDBP | Q13148 | 1.34365493 | 4.89034535 | 2.16715099 | 1.69E-05 | 0.00022102 |
| MDH1 | P40925 | 1.60487394 | 4.70800008 | 1.60532541 | 3.00E-05 | 0.00035396 |
| SOD1 | P00441 | 1.46028807 | 4.63786288 | 1.37508823 | 3.74E-05 | 0.00040115 |
| Oligo-SNCA | P37840 | 2.44499383 | 4.50274101 | 0.95950581 | 5.71E-05 | 0.00051792 |
| MAPT | P10636 | 0.61624078 | 4.50409266 | 0.9595037 | 5.67E-05 | 0.00051792 |
| TNF | P01375 | 0.56781759 | 4.4758619 | 0.89624658 | 6.19E-05 | 0.00052169 |
| ICAM1 | P05362 | 0.56131256 | 4.42733463 | 0.7468318 | 7.20E-05 | 0.00056619 |
| IGF1 | P05019 | 0.34230636 | 4.22819863 | 0.15888177 | 0.00013294 | 0.00098047 |
| AGRN | O00468 | 0.513112 | 4.08047405 | -0.3123496 | 0.00020869 | 0.00144857 |
| NPTX2 | P47972 | 0.60528757 | 4.04636934 | -0.4040851 | 0.00023179 | 0.0015195 |
| NRGN | Q92686 | 2.54037953 | 3.92970647 | -0.7253821 | 0.00032919 | 0.00204444 |
| FGF2 | P09038 | 1.69408488 | 3.89065246 | -0.8444424 | 0.00036992 | 0.00218253 |
| POSTN | Q15063 | 0.82918783 | 3.84679494 | -0.9831263 | 0.00042149 | 0.00230036 |
| PSEN1 | P49768 | 0.83938926 | 3.82595449 | -1.0380619 | 0.00044838 | 0.00230036 |
| PGF | P49763 | 0.57767099 | 3.82661236 | -1.0384938 | 0.0004475 | 0.00230036 |
| CX3CL1 | P78423 | 0.52129281 | 3.78694778 | -1.1633699 | 0.00050266 | 0.00247143 |
| GOT1 | P17174 | 0.69474391 | 3.76565388 | -1.2241009 | 0.00053585 | 0.00252919 |
| IGFBP7 | Q16270 | 0.38361108 | 3.74449803 | -1.2781509 | 0.00056924 | 0.00258348 |
| CHI3L1 | P36222 | 1.02262047 | 3.67636805 | -1.4649181 | 0.00069629 | 0.00304306 |
| BASP1 | P80723 | 0.50186094 | 3.6276597 | -1.5770859 | 0.00080165 | 0.00319449 |
| ACHE | P22303 | 0.60647878 | 3.63627378 | -1.5873411 | 0.00078259 | 0.00319449 |
| PDGFRB | P09619 | 0.67876864 | 3.62350903 | -1.6021394 | 0.00081216 | 0.00319449 |
| BACE1 | P56817 | 0.32452041 | 3.55460704 | -1.7897848 | 0.00098975 | 0.00376745 |
| IL12p70 | P29459\|P29460 | 0.55844791 | 3.42835557 | -2.11444 | 0.00142243 | 0.00508625 |
| REST | Q13127 | 0.55894363 | 3.42907099 | -2.152675 | 0.00141954 | 0.00508625 |
| PGK 1.00 | P00558 | 1.91371995 | 3.41125983 | -2.1907724 | 0.00149308 | 0.00518186 |
| IL33 | O95760 | 0.38226355 | 3.383871 | -2.2509632 | 0.00161117 | 0.00543193 |
| TAFA5 | Q7Z5A7 | 0.81044807 | 3.32868128 | -2.3875114 | 0.00188424 | 0.00600918 |
| CST3 | P01034 | 0.28639869 | 3.34059337 | -2.387983 | 0.00181999 | 0.00596554 |
| FLT1 | P17948 | 0.60414088 | 3.21734853 | -2.6967181 | 0.00256822 | 0.007975 |
| NCAM1 | P13591 | 0.31458329 | 3.19869271 | -2.7651242 | 0.00270074 | 0.00817147 |
| CCL3 | P10147 | 0.546836 | 3.0969937 | -3.0138508 | 0.00356993 | 0.01036976 |
| SLIT2 | O94813 | 0.41105196 | 3.09330951 | -3.0181801 | 0.00360305 | 0.01036976 |
| CCL2 | P13500 | 0.66196008 | 3.07268321 | -3.0863569 | 0.00381276 | 0.01071203 |
| IL6 | P05231 | 0.62265564 | 2.83754319 | -3.6512411 | 0.00711139 | 0.01951497 |
| IL18 | Q14116 | 0.93413516 | 2.79055791 | -3.7698621 | 0.00803066 | 0.02153677 |
| CCL22 | O00626 | 0.5090017 | 2.7538799 | -3.8605024 | 0.00882374 | 0.0231378 |
| HBA1 | P69905 | 1.29706484 | 2.73826852 | -3.8670661 | 0.00918284 | 0.02355598 |
| IL10 | P22301 | 0.62938766 | 2.65986985 | -4.0859984 | 0.01119959 | 0.02811813 |
| CSF2 | P04141 | 0.42688577 | 2.57067587 | -4.262276 | 0.01398587 | 0.03438192 |
| IL1B | P01584 | 0.2324924 | 2.53880983 | -4.3223871 | 0.01511826 | 0.0349795 |
| KDR | P35968 | 0.49129996 | 2.5425078 | -4.3334952 | 0.0149898 | 0.0349795 |

**Table S7**- Correlation of the same protein targets in plasma and serum (ranked by correlation coefficient).

| Target | Cor. Coeff. | *P* value | Lower ci | Upper ci | Adj. *P* value |
| --- | --- | --- | --- | --- | --- |
| NEFH | 0.97144179 | 2.40E-25 | 0.94629328 | 0.98490568 | 2.72E-23 |
| MME | 0.96556016 | 8.02E-24 | 0.93540197 | 0.98177133 | 4.53E-22 |
| KDR | 0.9592461 | 1.86E-22 | 0.92377325 | 0.97839669 | 7.00E-21 |
| SAA1 | 0.95599473 | 7.77E-22 | 0.91781055 | 0.97665496 | 2.19E-20 |
| IL13 | 0.95024577 | 7.61E-21 | 0.9073095 | 0.97356864 | 1.72E-19 |
| IFNG | 0.94816492 | 1.63E-20 | 0.90352179 | 0.97244943 | 3.07E-19 |
| MSLN | 0.93620077 | 7.58E-19 | 0.88187824 | 0.96599259 | 1.22E-17 |
| CRP | 0.92889562 | 5.58E-18 | 0.8687746 | 0.96203177 | 7.88E-17 |
| IL6 | 0.92267275 | 2.60E-17 | 0.85767832 | 0.95864672 | 3.26E-16 |
| NEFL | 0.92071287 | 4.11E-17 | 0.85419606 | 0.95757849 | 4.64E-16 |
| IL17A | 0.91524077 | 1.39E-16 | 0.84450487 | 0.95459056 | 1.43E-15 |
| TREM2 | 0.90609853 | 8.98E-16 | 0.8284165 | 0.94958091 | 8.22E-15 |
| IL5 | 0.90582892 | 9.46E-16 | 0.82794399 | 0.94943284 | 8.22E-15 |
| CRH | 0.90183908 | 2.01E-15 | 0.8209643 | 0.94723926 | 1.62E-14 |
| GFAP | 0.88771691 | 2.27E-14 | 0.79645216 | 0.93944073 | 1.71E-13 |
| CHI3L1 | 0.88009901 | 7.39E-14 | 0.78335298 | 0.93521161 | 5.22E-13 |
| CSF2 | 0.87887537 | 8.86E-14 | 0.78125688 | 0.93453083 | 5.89E-13 |
| ACHE | 0.82787623 | 4.41E-11 | 0.6958102 | 0.90579026 | 2.77E-10 |
| IL12p70 | 0.82483342 | 5.98E-11 | 0.69082762 | 0.90405252 | 3.56E-10 |
| VSNL1 | 0.81902663 | 1.05E-10 | 0.68135422 | 0.90072898 | 5.95E-10 |
| IL2 | 0.809582 | 2.53E-10 | 0.66604381 | 0.89530282 | 1.36E-09 |
| PDGFRB | 0.79996525 | 5.91E-10 | 0.65057768 | 0.8897515 | 3.03E-09 |
| TREM1 | 0.79227276 | 1.12E-09 | 0.63829481 | 0.88529177 | 5.53E-09 |
| FABP3 | 0.78351238 | 2.27E-09 | 0.62440163 | 0.88019201 | 1.07E-08 |
| PTN | 0.78195794 | 2.56E-09 | 0.6219469 | 0.87928477 | 1.16E-08 |
| CHIT1 | 0.78015689 | 7.72E-09 | 0.61347653 | 0.8802925 | 3.12E-08 |
| NGF | 0.77924952 | 3.16E-09 | 0.61767733 | 0.87770232 | 1.37E-08 |
| FOLR1 | 0.77339362 | 4.91E-09 | 0.60847851 | 0.87427356 | 2.06E-08 |
| CXCL10 | 0.75355909 | 2.00E-08 | 0.577647 | 0.8625847 | 7.80E-08 |
| GDNF | 0.75185307 | 2.24E-08 | 0.57501834 | 0.86157384 | 8.45E-08 |
| BACE1 | 0.74680821 | 3.13E-08 | 0.56726644 | 0.85857955 | 1.14E-07 |
| CCL4 | 0.73992512 | 4.88E-08 | 0.55674101 | 0.85448187 | 1.72E-07 |
| CCL22 | 0.7240932 | 1.29E-07 | 0.53275245 | 0.8450023 | 4.40E-07 |
| PDLIM5 | 0.71019799 | 2.85E-07 | 0.51194876 | 0.83661928 | 9.49E-07 |
| CCL3 | 0.70123551 | 4.66E-07 | 0.49865253 | 0.83118059 | 1.50E-06 |
| IL10 | 0.69972653 | 5.05E-07 | 0.49642323 | 0.83026245 | 1.59E-06 |
| CALB2 | 0.67150919 | 2.10E-06 | 0.4552243 | 0.81296202 | 6.42E-06 |
| IL33 | 0.66392696 | 3.01E-06 | 0.44430944 | 0.80827029 | 8.94E-06 |
| pTau217 | 0.66281686 | 3.16E-06 | 0.44271688 | 0.80758184 | 9.17E-06 |
| IL9 | 0.65910939 | 3.75E-06 | 0.43740813 | 0.80527972 | 1.06E-05 |
| FCN2 | 0.6503401 | 5.56E-06 | 0.42491251 | 0.79981692 | 1.53E-05 |
| AGRN | 0.63865018 | 9.23E-06 | 0.40838773 | 0.79249601 | 2.48E-05 |
| FGF2 | 0.62903039 | 1.38E-05 | 0.39490154 | 0.7864381 | 3.56E-05 |
| SFTPD | 0.62889711 | 1.39E-05 | 0.3947154 | 0.78635395 | 3.56E-05 |
| IL4 | 0.62551362 | 1.59E-05 | 0.38999636 | 0.78421588 | 3.99E-05 |
| CCL2 | 0.61275833 | 2.64E-05 | 0.37231679 | 0.77612162 | 6.44E-05 |
| ICAM1 | 0.61234827 | 2.68E-05 | 0.37175131 | 0.77586051 | 6.44E-05 |
| CD63 | 0.61079345 | 2.84E-05 | 0.36960878 | 0.77486995 | 6.56E-05 |
| PGF | 0.61076466 | 2.85E-05 | 0.36956913 | 0.7748516 | 6.56E-05 |
| NPTX2 | 0.60419853 | 3.65E-05 | 0.36054945 | 0.77065939 | 8.26E-05 |
| IL15 | 0.59881642 | 4.47E-05 | 0.35319006 | 0.76721235 | 9.89E-05 |
| CCL17 | 0.59775868 | 4.64E-05 | 0.35174729 | 0.76653376 | 0.00010091 |
| VCAM1 | 0.58901312 | 6.38E-05 | 0.33986285 | 0.7609086 | 0.00013598 |
| TIMP3 | 0.58429396 | 7.54E-05 | 0.33348284 | 0.75786247 | 0.0001578 |
| SLIT2 | 0.57770305 | 9.49E-05 | 0.3246106 | 0.75359544 | 0.00019492 |
| CXCL8 | 0.5541335 | 0.00020744 | 0.29324312 | 0.738214 | 0.00041858 |
| CCL26 | 0.52818164 | 0.00046002 | 0.25934288 | 0.72105351 | 0.00091196 |
| TNF | 0.51981434 | 0.00058678 | 0.24855233 | 0.71546979 | 0.00114321 |
| IGFBP7 | 0.51471128 | 0.0006786 | 0.24200425 | 0.71205207 | 0.0012997 |
| VEGFA | 0.50694645 | 0.00084299 | 0.23208812 | 0.70683365 | 0.00158763 |
| HBA1 | 0.49716858 | 0.00174533 | 0.2063988 | 0.70725449 | 0.00308159 |
| NPTX1 | 0.49463356 | 0.00117684 | 0.21648011 | 0.69851382 | 0.00218005 |
| S100B | 0.48740347 | 0.00142338 | 0.20738092 | 0.69360262 | 0.00259423 |
| CX3CL1 | 0.48469168 | 0.00152702 | 0.20398053 | 0.69175562 | 0.00273893 |
| IGF1 | 0.45916313 | 0.0028797 | 0.17229821 | 0.67423431 | 0.00500624 |
| SNAP25 | 0.45406177 | 0.00325038 | 0.16603752 | 0.67070377 | 0.00556504 |
| IL16 | 0.43993198 | 0.00450305 | 0.14881694 | 0.66087331 | 0.0075947 |
| CCL11 | 0.43659183 | 0.00485421 | 0.1447718 | 0.65853836 | 0.00806655 |
| SFRP1 | 0.4357697 | 0.0049442 | 0.14377764 | 0.65796299 | 0.00809702 |
| S100A12 | 0.42268503 | 0.00658407 | 0.12803395 | 0.64877054 | 0.01062858 |
| IL6R | 0.41126183 | 0.00838003 | 0.11440973 | 0.64069103 | 0.01333724 |
| PGK1 | 0.37350876 | 0.01759645 | 0.07016297 | 0.61362226 | 0.02761665 |
| AB40 | 0.37200266 | 0.01809462 | 0.06842223 | 0.61253055 | 0.02800948 |
| TEK | 0.36976904 | 0.01885521 | 0.06584404 | 0.61090982 | 0.02879242 |
| CNTN2 | 0.36777265 | 0.01955756 | 0.06354309 | 0.6094595 | 0.02946673 |
| CCL13 | 0.3635204 | 0.02112686 | 0.05865291 | 0.60636498 | 0.0314123 |
| GOT1 | 0.33565882 | 0.03422263 | 0.02696941 | 0.58590539 | 0.05022281 |
| NPY | 0.32696628 | 0.03946292 | 0.0172098 | 0.57945624 | 0.05717064 |
| NPTXR | 0.30425616 | 0.05628854 | -0.0080126 | 0.56245663 | 0.08051399 |
| CXCL1 | 0.28717095 | 0.07238445 | -0.0267295 | 0.54952202 | 0.10224303 |
| VEGFD | 0.28063726 | 0.0794194 | -0.0338295 | 0.54454212 | 0.11079497 |
| KLK6 | 0.27698242 | 0.08358104 | -0.0377873 | 0.54174831 | 0.11517875 |
| TAFA5 | 0.26712102 | 0.09565714 | -0.0484172 | 0.53418081 | 0.130232 |
| IL18 | 0.26220657 | 0.10215506 | -0.053688 | 0.53039348 | 0.13742288 |
| ENO2 | 0.25202003 | 0.11669121 | -0.0645575 | 0.52250898 | 0.15495072 |
| RUVBL2 | 0.2511992 | 0.1179271 | -0.0654301 | 0.52187162 | 0.15495072 |
| FLT1 | 0.24569893 | 0.12646422 | -0.0712649 | 0.517593 | 0.16425812 |
| PARK7 | 0.23858344 | 0.13818309 | -0.078781 | 0.51203765 | 0.17743965 |
| CST3 | 0.19082174 | 0.23821146 | -0.128315 | 0.47414652 | 0.29908772 |
| pTau231 | 0.1872458 | 0.24728784 | -0.1319607 | 0.47126672 | 0.30707171 |
| POSTN | 0.17635999 | 0.27633493 | -0.1430058 | 0.46246267 | 0.33412186 |
| AB38 | 0.17213395 | 0.28818942 | -0.1472723 | 0.45902952 | 0.34279374 |
| MAPT | 0.16136772 | 0.31985523 | -0.1580883 | 0.45024438 | 0.37649626 |
| ANXA5 | 0.15460985 | 0.34080506 | -0.1648385 | 0.44470131 | 0.39702033 |
| MDH1 | 0.14135384 | 0.38428306 | -0.1779931 | 0.43376327 | 0.4431019 |
| IL1B | 0.12170762 | 0.45438432 | -0.1972813 | 0.41739235 | 0.51864069 |
| AB42 | 0.1007191 | 0.53632501 | -0.217618 | 0.39968825 | 0.60604726 |
| SQSTM1 | 0.06304686 | 0.69913819 | -0.25344 | 0.36734173 | 0.75964053 |
| SOD1 | 0.05546934 | 0.73388939 | -0.260542 | 0.36074529 | 0.78235378 |
| SNCA | 0.02428046 | 0.8817782 | -0.2894179 | 0.3332691 | 0.9312237 |
| pTau181 | 0.01962624 | 0.90432262 | -0.2936786 | 0.32912341 | 0.94374293 |
| UCHL1 | 0.01838751 | 0.9103361 | -0.2948105 | 0.32801801 | 0.94374293 |
| pTDP43 | 0.00504688 | 0.9753435 | -0.306945 | 0.31605933 | 0.98648385 |
| OligoSNCA | 0.0042233 | 0.97936615 | -0.3076908 | 0.31531782 | 0.98648385 |
| REST | -0.0027663 | 0.98648385 | -0.3140051 | 0.30900934 | 0.98648385 |
| BASP1 | -0.0107232 | 0.94763982 | -0.3211598 | 0.30179423 | 0.97348455 |
| TARDBP | -0.0583578 | 0.7205766 | -0.3632634 | 0.25783883 | 0.77547768 |
| IL7 | -0.0694144 | 0.67039354 | -0.3728613 | 0.24744554 | 0.73548029 |
| PRDX6 | -0.0755482 | 0.64313459 | -0.378158 | 0.24164809 | 0.71249224 |
| CD40LG | -0.079988 | 0.62368523 | -0.3819796 | 0.23743759 | 0.69778644 |
| YWHAZ | -0.1757802 | 0.27794208 | -0.4619922 | 0.14359182 | 0.33412186 |
| PSEN1 | -0.1764452 | 0.2760993 | -0.4625318 | 0.14291965 | 0.33412186 |
| NRGN | -0.2273679 | 0.15825663 | -0.5032345 | 0.09055524 | 0.20093257 |

**Table S8**- Correlation of the same protein targets in CSF and plasma (ranked by correlation coefficient).

| Target | Cor. Coeff. | *P* value | Lower CI | Upper CI | Adj. *P* value |
| --- | --- | --- | --- | --- | --- |
| PDGFRB | 0.72839218 | 9.95E-08 | 0.539236 | 0.84758391 | 5.77E-06 |
| pTau217 | 0.72134391 | 1.51E-07 | 0.52861782 | 0.84334835 | 5.84E-06 |
| NEFL | 0.70037798 | 4.88E-07 | 0.49738531 | 0.83065891 | 1.42E-05 |
| IL13 | 0.67194423 | 2.06E-06 | 0.45585252 | 0.81323065 | 4.63E-05 |
| CCL4 | 0.66879124 | 2.39E-06 | 0.45130423 | 0.81128231 | 4.63E-05 |
| ACHE | 0.64338965 | 7.54E-06 | 0.41506927 | 0.7954695 | 0.0001249 |
| CCL17 | 0.61175965 | 2.74E-05 | 0.37093989 | 0.7754856 | 0.00039716 |
| KDR | 0.59565952 | 7.97E-05 | 0.34088909 | 0.76892273 | 0.00102691 |
| CHIT1 | 0.58616974 | 0.00011019 | 0.32793833 | 0.76289516 | 0.00116202 |
| TREM2 | 0.57899511 | 9.07E-05 | 0.32634638 | 0.7544331 | 0.0010525 |
| CCL13 | 0.5450697 | 0.00027599 | 0.2813283 | 0.73224758 | 0.00266794 |
| CSF2 | 0.53998036 | 0.00032285 | 0.27467356 | 0.72888478 | 0.00288079 |
| SAA1 | 0.50627298 | 0.00138824 | 0.21801813 | 0.71328348 | 0.01150259 |
| MSLN | 0.45132977 | 0.00346554 | 0.16269415 | 0.66880896 | 0.02680016 |
| pTau231 | 0.44030954 | 0.00446479 | 0.14927481 | 0.66113699 | 0.0323697 |
| SFTPD | 0.42655305 | 0.00605646 | 0.13267261 | 0.65149485 | 0.04132643 |
| ICAM1 | 0.41964116 | 0.00702671 | 0.12439269 | 0.64662261 | 0.04528327 |
| IL1B | 0.39822346 | 0.01092907 | 0.0989944 | 0.63140661 | 0.06338858 |
| VSNL1 | 0.39244988 | 0.01225348 | 0.09221384 | 0.62727386 | 0.06768589 |
| CCL22 | 0.35918836 | 0.02283249 | 0.05368596 | 0.6032048 | 0.1203895 |
| CCL3 | 0.34997819 | 0.02684222 | 0.04317595 | 0.59646053 | 0.13537815 |
| CXCL8 | 0.34616704 | 0.028664 | 0.0388467 | 0.59365955 | 0.13854267 |
| IL18 | 0.34265267 | 0.03043333 | 0.03486477 | 0.59107136 | 0.1404541 |
| CCL11 | 0.34065025 | 0.03148109 | 0.03260031 | 0.58959436 | 0.1404541 |
| SFRP1 | 0.32153308 | 0.04305798 | 0.01113946 | 0.57540913 | 0.17838304 |
| IL6R | 0.31469804 | 0.04795315 | 0.00353526 | 0.57030009 | 0.18802138 |
| KLK6 | 0.31380289 | 0.04862622 | 0.00254203 | 0.56962951 | 0.18802138 |
| GDNF | 0.28337528 | 0.07640905 | -0.030858 | 0.54663128 | 0.28591774 |
| IL33 | 0.27819558 | 0.08218128 | -0.0364747 | 0.54267631 | 0.29790715 |
| IGFBP7 | 0.26660926 | 0.09631859 | -0.0489669 | 0.53378692 | 0.33857444 |
| IL12p70 | 0.26202111 | 0.10240671 | -0.0538866 | 0.53025035 | 0.34938761 |
| IL9 | 0.25712046 | 0.10922953 | -0.0591245 | 0.52646256 | 0.35977545 |
| CCL26 | 0.25494988 | 0.11235942 | -0.0614389 | 0.52478147 | 0.35977545 |
| CST3 | 0.24596487 | 0.12604111 | -0.0709832 | 0.51780018 | 0.38475706 |
| BDNF | 0.22986365 | 0.3747926 | -0.2819295 | 0.6398194 | 0.65872638 |
| SNCA | 0.22582065 | 0.16118401 | -0.0921726 | 0.50201559 | 0.47722145 |
| IL10 | 0.22276175 | 0.16708668 | -0.0953652 | 0.4996025 | 0.47722145 |
| ANXA5 | 0.21727081 | 0.17807062 | -0.1010798 | 0.49526002 | 0.48037655 |
| NGF | 0.20990132 | 0.19360978 | -0.1087166 | 0.48940999 | 0.51042578 |
| VEGFD | 0.20436873 | 0.20588716 | -0.1144252 | 0.48500154 | 0.52687424 |
| SOD1 | 0.20303211 | 0.20893289 | -0.1158012 | 0.48393435 | 0.52687424 |
| GOT1 | 0.19644955 | 0.22438928 | -0.12256 | 0.47866648 | 0.55381183 |
| SQSTM1 | 0.18999842 | 0.24028092 | -0.1291551 | 0.47348401 | 0.58067888 |
| IL6 | 0.18659595 | 0.2489619 | -0.1326222 | 0.47074273 | 0.5893792 |
| IL7 | 0.17741645 | 0.27342205 | -0.1419373 | 0.46331957 | 0.61226921 |
| NEFH | 0.17674719 | 0.275265 | -0.1426143 | 0.46277679 | 0.61226921 |
| PRDX6 | 0.17117739 | 0.29091765 | -0.1482364 | 0.45825125 | 0.61226921 |
| CXCL10 | 0.16955663 | 0.29557824 | -0.1498686 | 0.45693155 | 0.61226921 |
| FCN2 | 0.16542297 | 0.39112691 | -0.2140609 | 0.50152213 | 0.66059453 |
| PSEN1 | 0.16436428 | 0.31083036 | -0.1550856 | 0.45269517 | 0.62848144 |
| SLIT2 | 0.16322555 | 0.31424072 | -0.1562274 | 0.45176436 | 0.62848144 |
| IFNG | 0.1572036 | 0.33266665 | -0.1622513 | 0.44683144 | 0.65017623 |
| NRGN | 0.15299089 | 0.3459464 | -0.1664512 | 0.44337006 | 0.65017623 |
| NPTX2 | 0.14972724 | 0.35645397 | -0.169697 | 0.44068252 | 0.65017623 |
| TREM1 | 0.14814639 | 0.36161229 | -0.1712667 | 0.43937885 | 0.65017623 |
| FLT1 | 0.14732112 | 0.36432289 | -0.1720856 | 0.43869779 | 0.65017623 |
| RUVBL2 | 0.14100831 | 0.38545815 | -0.1783344 | 0.43347701 | 0.66059453 |
| VEGFA | 0.13172829 | 0.41779978 | -0.187474 | 0.42576649 | 0.69235392 |
| IL16 | 0.12823032 | 0.43037715 | -0.1909047 | 0.42284899 | 0.69338541 |
| PDLIM5 | 0.12310389 | 0.44918523 | -0.1959185 | 0.41856221 | 0.71377379 |
| GFAP | 0.11591049 | 0.47631462 | -0.2029259 | 0.41252473 | 0.72700652 |
| S100A12 | 0.11315023 | 0.48694876 | -0.2056062 | 0.41020108 | 0.73358515 |
| ENO2 | 0.1058797 | 0.51553764 | -0.2126432 | 0.40406206 | 0.74761097 |
| NCAM1 | 0.10315529 | 0.52646158 | -0.2152716 | 0.40175472 | 0.74761097 |
| BACE1 | 0.09917464 | 0.54262392 | -0.2191037 | 0.39837661 | 0.74761097 |
| TNF | 0.09819959 | 0.54661875 | -0.2200408 | 0.39754791 | 0.74761097 |
| CXCL1 | 0.09634498 | 0.55425547 | -0.2218218 | 0.39597032 | 0.74761097 |
| pTau181 | 0.09319583 | 0.56733631 | -0.224841 | 0.39328751 | 0.75644841 |
| AB42 | 0.0904381 | 0.57890651 | -0.2274799 | 0.39093394 | 0.76310403 |
| FOLR1 | 0.08263489 | 0.6122076 | -0.2349217 | 0.38425297 | 0.79793349 |
| NPY | 0.07762713 | 0.63399719 | -0.239678 | 0.37994872 | 0.79938776 |
| CRH | 0.06880548 | 0.67312312 | -0.2480198 | 0.37233438 | 0.80722862 |
| MDH1 | 0.06686372 | 0.6818551 | -0.2498497 | 0.37065286 | 0.80722862 |
| CX3CL1 | 0.05701381 | 0.72676106 | -0.2590972 | 0.36209228 | 0.85155842 |
| CD63 | 0.05210063 | 0.74951312 | -0.2636883 | 0.3578029 | 0.85159758 |
| TEK | 0.05067401 | 0.75615992 | -0.2650188 | 0.35655499 | 0.85159758 |
| POSTN | 0.04347387 | 0.78996196 | -0.2717152 | 0.35024008 | 0.87271988 |
| CCL2 | 0.03359538 | 0.83695011 | -0.2808532 | 0.34153052 | 0.90533839 |
| OligoSNCA | 0.03235335 | 0.84290126 | -0.2819981 | 0.3404317 | 0.90533839 |
| IL2 | 0.01868695 | 0.908882 | -0.2945369 | 0.3282853 | 0.9248273 |
| IL5 | 0.00613323 | 0.97003831 | -0.3059607 | 0.31703686 | 0.97847343 |
| CHI3L1 | 0.0011248 | 0.99450402 | -0.3104933 | 0.31252464 | 0.99450402 |
| IL17A | -0.0226057 | 0.88988106 | -0.3317787 | 0.29095254 | 0.91350622 |
| CALB2 | -0.0243567 | 0.88140969 | -0.3333369 | 0.28934805 | 0.9128886 |
| IGF1 | -0.0263092 | 0.8719791 | -0.3350724 | 0.28755688 | 0.91125744 |
| PGF | -0.0267973 | 0.86962409 | -0.335506 | 0.28710874 | 0.91125744 |
| SNAP25 | -0.0287379 | 0.86027246 | -0.3372284 | 0.2853257 | 0.91125744 |
| YWHAZ | -0.0329417 | 0.84008118 | -0.3409523 | 0.28145586 | 0.90533839 |
| MAPT | -0.0474761 | 0.77112196 | -0.3537537 | 0.26799675 | 0.86009757 |
| PGK1 | -0.0507806 | 0.75566254 | -0.3566483 | 0.26491938 | 0.85159758 |
| PARK7 | -0.0528689 | 0.74594085 | -0.3584745 | 0.26297133 | 0.85159758 |
| S100B | -0.0668385 | 0.681969 | -0.370631 | 0.24987346 | 0.80722862 |
| IL15 | -0.0702249 | 0.66676669 | -0.3735623 | 0.24668078 | 0.80722862 |
| CNTN2 | -0.0712162 | 0.66234091 | -0.3744192 | 0.24574488 | 0.80722862 |
| REST | -0.0760107 | 0.64109718 | -0.3785566 | 0.24121002 | 0.7996481 |
| AGRN | -0.0777334 | 0.63353149 | -0.3800402 | 0.23957723 | 0.79938776 |
| YWHAG | -0.0797638 | 0.62466141 | -0.3817868 | 0.23765048 | 0.79938776 |
| TIMP3 | -0.1004535 | 0.53740568 | -0.3994628 | 0.21787359 | 0.74761097 |
| PTN | -0.1016912 | 0.53237857 | -0.4005132 | 0.21668215 | 0.74761097 |
| UCHL1 | -0.1054309 | 0.51732943 | -0.4036822 | 0.21307647 | 0.74761097 |
| HBA1 | -0.108552 | 0.55426331 | -0.4405698 | 0.24958927 | 0.74761097 |
| NPTXR | -0.1172574 | 0.47117024 | -0.4136572 | 0.20161634 | 0.72700652 |
| MME | -0.1200137 | 0.46073539 | -0.4159717 | 0.19893287 | 0.72223385 |
| FGF2 | -0.1294474 | 0.42597737 | -0.4238648 | 0.18971194 | 0.69338541 |
| CD40LG | -0.1446702 | 0.39293985 | -0.4477027 | 0.1881694 | 0.66059453 |
| VCAM1 | -0.1480879 | 0.3618039 | -0.4393306 | 0.17132476 | 0.65017623 |
| AB40 | -0.1497321 | 0.35643805 | -0.4406866 | 0.16969215 | 0.65017623 |
| IL4 | -0.1710268 | 0.29134857 | -0.4581287 | 0.14838811 | 0.61226921 |
| NPTX1 | -0.1744857 | 0.28155249 | -0.4609411 | 0.1448995 | 0.61226921 |
| pTDP43 | -0.1817864 | 0.26158793 | -0.4668584 | 0.13750988 | 0.60688399 |
| AB38 | -0.2198808 | 0.17278708 | -0.4973258 | 0.09836614 | 0.47722145 |
| BASP1 | -0.2200334 | 0.1724817 | -0.4974465 | 0.09820734 | 0.47722145 |
| FABP3 | -0.2533186 | 0.11475596 | -0.5235167 | 0.06317601 | 0.35977545 |
| TAFA5 | -0.3256021 | 0.04034174 | -0.5784412 | -0.0156835 | 0.17332006 |
| TARDBP | -0.3986953 | 0.01082645 | -0.6317438 | -0.0995497 | 0.06338858 |
| CRP | -0.8661632 | 5.26E-13 | -0.9274342 | -0.7596104 | 6.10E-11 |

**Table S9**- Correlation of the same protein targets in CSF and serum (ranked by correlation coefficient).

| Target | Cor. Coeff. | *P* value | Lower CI | Upper CI | Adj. *P* value |
| --- | --- | --- | --- | --- | --- |
| NEFL | 0.68192383 | 1.27E-06 | 0.47032319 | 0.81937658 | 5.64E-05 |
| ACHE | 0.67850484 | 1.50E-06 | 0.46535272 | 0.81727455 | 5.64E-05 |
| IL13 | 0.66563546 | 2.78E-06 | 0.44676321 | 0.80932908 | 7.84E-05 |
| PDGFRB | 0.63481362 | 1.08E-05 | 0.4029971 | 0.79008363 | 0.00024516 |
| KDR | 0.6216126 | 3.11E-05 | 0.37680829 | 0.78525142 | 0.00058541 |
| CCL4 | 0.59384732 | 5.36E-05 | 0.34642228 | 0.76402117 | 0.00086489 |
| CCL17 | 0.55233458 | 0.00021968 | 0.29087188 | 0.73703211 | 0.00310294 |
| pTau217 | 0.53994779 | 0.00032317 | 0.27463105 | 0.72886323 | 0.00405757 |
| SAA1 | 0.52331151 | 0.00088898 | 0.23998508 | 0.72448479 | 0.01004543 |
| CSF2 | 0.49123085 | 0.00128771 | 0.21219171 | 0.69620484 | 0.01322831 |
| TREM2 | 0.47249991 | 0.00207988 | 0.18877617 | 0.6834182 | 0.0195855 |
| CCL22 | 0.45408809 | 0.00324836 | 0.16606976 | 0.67072201 | 0.02823575 |
| CCL13 | 0.39135413 | 0.01251966 | 0.09093011 | 0.62648803 | 0.10105153 |
| MSLN | 0.37873346 | 0.01595678 | 0.076216 | 0.61740229 | 0.12020772 |
| CHIT1 | 0.37640732 | 0.0198477 | 0.06448332 | 0.62132582 | 0.14017435 |
| ICAM1 | 0.32442981 | 0.04110957 | 0.01437304 | 0.57756841 | 0.25807674 |
| IL12p70 | 0.31117868 | 0.05064334 | -0.0003662 | 0.56766171 | 0.28613488 |
| SFRP1 | 0.30368316 | 0.05677748 | -0.0086439 | 0.56202487 | 0.30551692 |
| REST | 0.29223871 | 0.06727287 | -0.0212006 | 0.55337181 | 0.33051453 |
| pTau231 | 0.25437922 | 0.11319346 | -0.0620468 | 0.52433915 | 0.45723328 |
| IL33 | 0.25240574 | 0.11611386 | -0.0641473 | 0.52280837 | 0.45723328 |
| S100B | 0.23957603 | 0.13650183 | -0.0777347 | 0.51281398 | 0.51415688 |
| NGF | 0.23642786 | 0.14188692 | -0.0810508 | 0.51035018 | 0.51720069 |
| NEFH | 0.22111203 | 0.17033405 | -0.0970843 | 0.49829929 | 0.58445927 |
| CCL3 | 0.21835844 | 0.175855 | -0.0999495 | 0.49612127 | 0.58445927 |
| GFAP | 0.20293964 | 0.20914474 | -0.1158964 | 0.4838605 | 0.62524912 |
| FOLR1 | 0.20245361 | 0.21026077 | -0.1163964 | 0.48347221 | 0.62524912 |
| IGFBP7 | 0.19871957 | 0.21897295 | -0.1202325 | 0.48048543 | 0.63446009 |
| IL10 | 0.19502446 | 0.22783615 | -0.1240193 | 0.47752334 | 0.64363712 |
| GDNF | 0.18837789 | 0.2443896 | -0.1308074 | 0.47217908 | 0.6648818 |
| POSTN | 0.18153362 | 0.26226307 | -0.1377663 | 0.46665396 | 0.6648818 |
| IL18 | 0.17865017 | 0.27004605 | -0.1406887 | 0.46431958 | 0.6648818 |
| CCL2 | 0.17666191 | 0.27550041 | -0.1427005 | 0.46270761 | 0.6648818 |
| CNTN2 | 0.17410002 | 0.28263406 | -0.1452889 | 0.46062778 | 0.6648818 |
| SFTPD | 0.16971396 | 0.29512373 | -0.1497102 | 0.45705971 | 0.6648818 |
| AB38 | 0.16800682 | 0.3000794 | -0.1514276 | 0.45566845 | 0.6648818 |
| VSNL1 | 0.1634048 | 0.31370232 | -0.1560477 | 0.45191093 | 0.66639117 |
| IFNG | 0.16182997 | 0.31845242 | -0.1576255 | 0.45062273 | 0.66639117 |
| IL6 | 0.15701231 | 0.33326272 | -0.1624422 | 0.44667445 | 0.67406561 |
| CALB2 | 0.15307312 | 0.34568413 | -0.1663693 | 0.44343771 | 0.67406561 |
| OligoSNCA | 0.14946291 | 0.35731336 | -0.1699596 | 0.44046463 | 0.67406561 |
| IL6R | 0.14936496 | 0.35763214 | -0.1700569 | 0.44038387 | 0.67406561 |
| IL17A | 0.14927934 | 0.35791094 | -0.1701419 | 0.44031328 | 0.67406561 |
| BACE1 | 0.14699985 | 0.3653814 | -0.1724042 | 0.43843257 | 0.67685406 |
| CCL11 | 0.12666065 | 0.43608887 | -0.1924417 | 0.42153782 | 0.74706096 |
| TNF | 0.12565526 | 0.43976922 | -0.1934253 | 0.42069735 | 0.74706096 |
| pTau181 | 0.12517873 | 0.44151961 | -0.1938913 | 0.4202988 | 0.74706096 |
| TARDBP | 0.12333611 | 0.44832368 | -0.1956918 | 0.41875668 | 0.74706096 |
| CRH | 0.12300327 | 0.44955881 | -0.1960168 | 0.41847793 | 0.74706096 |
| NPTX1 | 0.11606813 | 0.47571101 | -0.2027727 | 0.41265732 | 0.75711753 |
| IL1B | 0.10223663 | 0.53017048 | -0.2161568 | 0.40097583 | 0.7987902 |
| AB40 | 0.09668844 | 0.55283743 | -0.2214921 | 0.39626261 | 0.80090551 |
| TEK | 0.0912221 | 0.57560639 | -0.2267301 | 0.39160344 | 0.82333572 |
| IL15 | 0.07957008 | 0.62550555 | -0.2378344 | 0.38162027 | 0.86197715 |
| IL16 | 0.07678925 | 0.63767351 | -0.2404723 | 0.37922727 | 0.86815791 |
| CST3 | 0.06193518 | 0.70420071 | -0.254484 | 0.36637589 | 0.90538699 |
| CCL26 | 0.05559955 | 0.73328756 | -0.2604202 | 0.3608589 | 0.91923667 |
| MAPT | 0.05194379 | 0.75024304 | -0.2638347 | 0.35766575 | 0.91923667 |
| PDLIM5 | 0.05075696 | 0.75577296 | -0.2649414 | 0.35662758 | 0.91923667 |
| CX3CL1 | 0.04993238 | 0.75962216 | -0.2657099 | 0.35590583 | 0.91923667 |
| TREM1 | 0.04991359 | 0.75970994 | -0.2657274 | 0.35588938 | 0.91923667 |
| NPTX2 | 0.0463541 | 0.77639097 | -0.2690401 | 0.35276955 | 0.91923667 |
| KLK6 | 0.04628845 | 0.77669955 | -0.2691011 | 0.35271195 | 0.91923667 |
| IL9 | 0.04281944 | 0.7930542 | -0.2723223 | 0.34966473 | 0.91923667 |
| CXCL10 | 0.03666505 | 0.82228086 | -0.2780197 | 0.34424263 | 0.92228022 |
| SQSTM1 | 0.02894455 | 0.85927783 | -0.2851357 | 0.33741163 | 0.94270287 |
| VEGFD | 0.01954631 | 0.90471052 | -0.2937517 | 0.32905211 | 0.96006429 |
| PTN | 0.01305241 | 0.93628791 | -0.2996754 | 0.32324745 | 0.97475237 |
| PGF | 0.01156865 | 0.94351813 | -0.3010255 | 0.32191789 | 0.97475237 |
| NPTXR | 0.00793134 | 0.96126018 | -0.3043299 | 0.3186534 | 0.97475237 |
| CHI3L1 | 0.00693448 | 0.96612624 | -0.3052342 | 0.31775742 | 0.97475237 |
| NPY | -0.002645 | 0.98707634 | -0.3138957 | 0.30911903 | 0.98707634 |
| AB42 | -0.009068 | 0.9557133 | -0.3196744 | 0.30329807 | 0.97475237 |
| IL7 | -0.0186446 | 0.90908742 | -0.3282475 | 0.29457559 | 0.96006429 |
| pTDP43 | -0.023328 | 0.8863849 | -0.3324217 | 0.29029086 | 0.95391898 |
| VEGFA | -0.0254674 | 0.87604256 | -0.3343245 | 0.28832932 | 0.95185394 |
| AGRN | -0.0289643 | 0.85918268 | -0.3374292 | 0.28511755 | 0.94270287 |
| NRGN | -0.0362337 | 0.82433896 | -0.3438618 | 0.27841824 | 0.92228022 |
| IL2 | -0.0404873 | 0.80409898 | -0.3476125 | 0.27448392 | 0.91923667 |
| CXCL8 | -0.0415015 | 0.7992911 | -0.3485053 | 0.27354428 | 0.91923667 |
| FCN2 | -0.0478392 | 0.80534894 | -0.4072052 | 0.32435351 | 0.91923667 |
| ENO2 | -0.0617423 | 0.70508013 | -0.3662083 | 0.25466506 | 0.90538699 |
| IL5 | -0.0682256 | 0.67572644 | -0.3718324 | 0.24856651 | 0.88787311 |
| S100A12 | -0.0704741 | 0.66565279 | -0.3737778 | 0.24644551 | 0.88492666 |
| MME | -0.0739141 | 0.65035357 | -0.3767488 | 0.2431948 | 0.8748804 |
| CD40LG | -0.0847718 | 0.61788649 | -0.3978624 | 0.24600436 | 0.86197715 |
| PSEN1 | -0.0878906 | 0.58968827 | -0.3887563 | 0.2299135 | 0.83293468 |
| PRDX6 | -0.0969853 | 0.55161328 | -0.3965152 | 0.22120715 | 0.80090551 |
| SNAP25 | -0.100223 | 0.53834452 | -0.3992671 | 0.21809539 | 0.8004333 |
| SNCA | -0.10671 | 0.51223107 | -0.4047645 | 0.21184121 | 0.78219068 |
| VCAM1 | -0.1084684 | 0.50526343 | -0.406251 | 0.21014137 | 0.7821201 |
| CD63 | -0.1117176 | 0.49251619 | -0.4089935 | 0.2069954 | 0.7729768 |
| SOD1 | -0.1174596 | 0.4704005 | -0.4138271 | 0.20141964 | 0.75711753 |
| SLIT2 | -0.120725 | 0.45806278 | -0.4165684 | 0.19823958 | 0.75016078 |
| UCHL1 | -0.1328056 | 0.41396834 | -0.4266638 | 0.18641576 | 0.74251465 |
| RUVBL2 | -0.1350276 | 0.40612963 | -0.4285127 | 0.18423093 | 0.740204 |
| TAFA5 | -0.1559949 | 0.33644421 | -0.4458392 | 0.16345752 | 0.67406561 |
| IL4 | -0.1657926 | 0.30658581 | -0.4538619 | 0.15365222 | 0.66623455 |
| ANXA5 | -0.1681417 | 0.29968596 | -0.4557784 | 0.15129198 | 0.6648818 |
| FLT1 | -0.1706335 | 0.29247636 | -0.4578085 | 0.14878434 | 0.6648818 |
| FGF2 | -0.1791054 | 0.26880736 | -0.4646884 | 0.1402277 | 0.6648818 |
| TIMP3 | -0.179961 | 0.26648924 | -0.4653813 | 0.13936086 | 0.6648818 |
| BASP1 | -0.2150175 | 0.18272411 | -0.493474 | 0.10341884 | 0.58993785 |
| FABP3 | -0.220269 | 0.17201089 | -0.4976329 | 0.09796209 | 0.58445927 |
| HBA1 | -0.2484125 | 0.19381406 | -0.563605 | 0.12992181 | 0.6083608 |
| PARK7 | -0.2515863 | 0.11734305 | -0.5221722 | 0.06501867 | 0.45723328 |
| CXCL1 | -0.2534373 | 0.11458029 | -0.5236088 | 0.06304967 | 0.45723328 |
| GOT1 | -0.2699765 | 0.09203026 | -0.5363765 | 0.04534648 | 0.41597679 |
| YWHAZ | -0.2799854 | 0.08014952 | -0.5440443 | 0.03453609 | 0.37737066 |
| IGF1 | -0.2957694 | 0.06388339 | -0.5560473 | 0.01733734 | 0.32812831 |
| PGK1 | -0.3207566 | 0.04359277 | -0.5748297 | -0.0102738 | 0.25926224 |
| MDH1 | -0.3508472 | 0.02644056 | -0.5970983 | -0.0441647 | 0.17575197 |
| CRP | -0.8875516 | 2.34E-14 | -0.9393491 | -0.7961669 | 2.64E-12 |

**Figure S1.** Plasma proteins passing multiple testing correction in biologically determined AD compared to non-AD (Cohort 1).

**Figure S2.** Nominally significant plasma proteins in biologically determined AD compared to non-AD (Cohort 1).

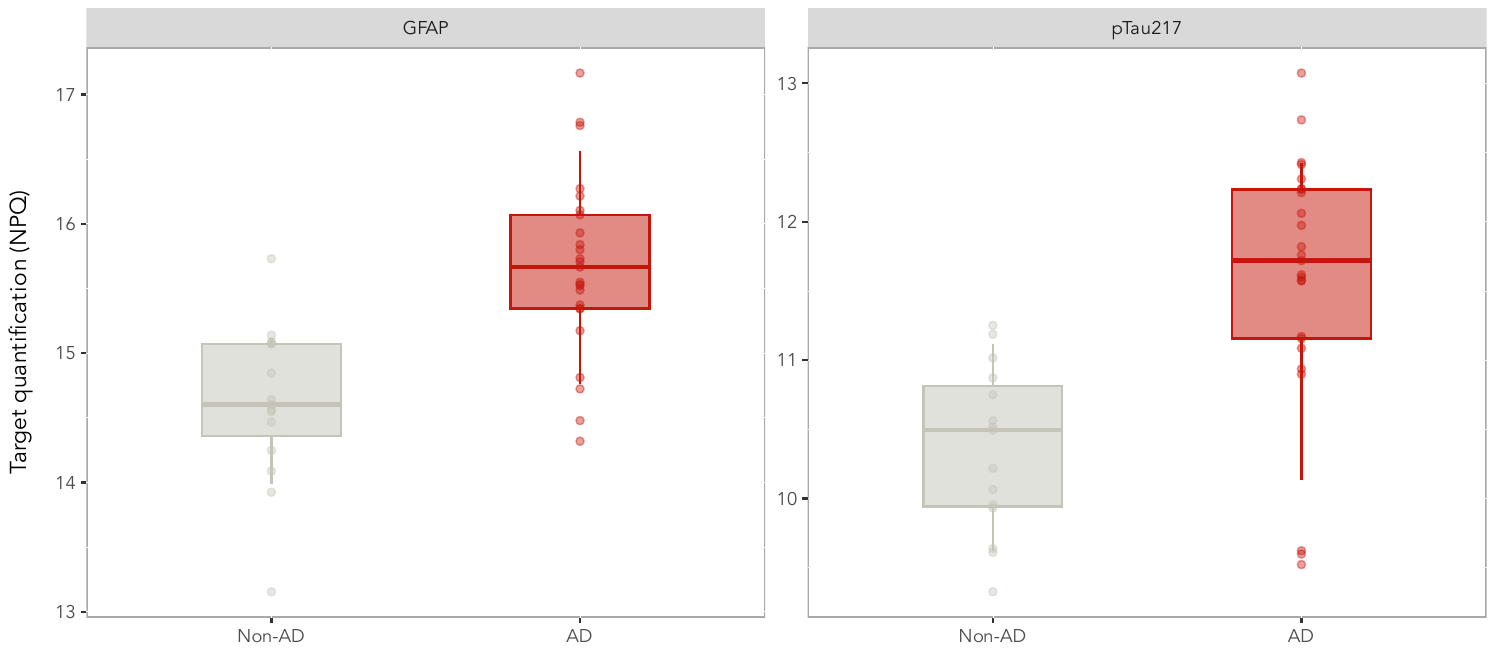

**Figure S3.** Serum proteins passing multiple testing correction in biologically determined AD compared to non-AD (Cohort 1).

**Figure S4.** Nominally significant serum proteins in biologically determined AD compared to non-AD (Cohort 1).

**Figure S5.** CSF proteins passing multiple testing correction in biologically determined AD compared to non-AD (cohort 1).

**Figure S6**. Nominally significant CSF proteins in biologically determined AD compared to non-AD (cohort 1).

**Figure S7**. Comparison between NULISAseq plasma pTau217 and other assays.

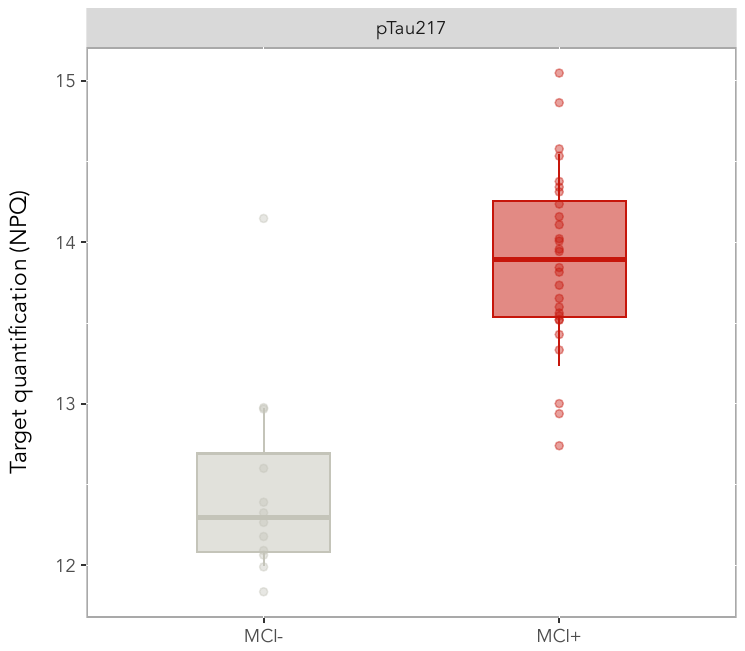

**Figure S8.** Plasma proteins passing multiple testing correction in MCI Aβ+ compared to MCI Aβ- (Cohort 2).

**Figure S9.** Nominally significant plasma proteins in MCI Aβ+ compared to MCI Aβ-.

**Figure S10.** Plasma proteins passing multiple testing correction in LB+ compared to AD.

**Figure S11.** Nominally significant plasma proteins in LB+ compared to AD.

**Figure S12.** Plasma proteins of interest compared between LB+ and AD.

**Figure S13.** Plasma proteins passing multiple testing correction in *GRN*+ compared to *GRN*-.

**Figure S14.** Plasma proteins of interest compared between *GRN*+ and *GRN*-.

**Figure S15** - Correlation of the same protein targets in plasma and serum (ranked by correlation coefficient). The bars indicate the confidence intervals (CI) at 95%.

**Figure S16** - Correlation of the same protein targets in CSF and plasma (ranked by correlation coefficient). The bars indicate the confidence intervals (CI) at 95%.

**Figure S17** - Correlation of the same protein targets in CSF and serum (ranked by correlation coefficient). The bars indicate the confidence intervals (CI) at 95%.
